# Multimorbidity Profiles and Severe In-Hospital Outcomes in Adults with Respiratory Syncytial Virus

**DOI:** 10.1101/2025.05.05.25326992

**Authors:** Kevin C. Ma, Diya Surie, Yuwei Zhu, Carlos G. Grijalva, Paul W. Blair, Basmah Safdar, Adit A. Ginde, Ithan D. Peltan, Samuel M. Brown, Manjusha Gaglani, Shekhar Ghamande, Cristie Columbus, Nicholas M. Mohr, Kevin W. Gibbs, David N. Hager, Matthew E. Prekker, Michelle N. Gong, Amira Mohamed, Nicholas J. Johnson, Jay S. Steingrub, Akram Khan, Catherine L. Hough, Abhijit Duggal, Alexandra June Gordon, Nida Qadir, Steven Y. Chang, Christopher Mallow, Laurence W. Busse, Jennie H. Kwon, Matthew C. Exline, Ivana A. Vaughn, Mayur Ramesh, Adam S. Lauring, Emily T. Martin, Aleda M. Leis, Jarrod M. Mosier, Estelle S. Harris, Adrienne Baughman, Cassandra Johnson, Jonathan D. Casey, Natasha Halasa, James D. Chappell, Nathaniel Lewis, Sascha Ellington, Wesley H. Self, Fatimah S. Dawood, the Investigating Respiratory Viruses in the Acutely Ill (IVY) Network

## Abstract

**Background:** Adults hospitalized with acute respiratory infections, including respiratory syncytial virus (RSV), often have multiple underlying conditions. Few data are available on the combined effect of conditions on risk of severe outcomes from RSV disease.

**Methods:** We enrolled adults hospitalized with RSV at 26 hospitals in 20 US states admitted January 2022–July 2024. Seventeen underlying conditions were selected after excluding those with rare prevalence (≤1%) or high pairwise correlation (≥0.7). We applied Bayesian profile regression to identify profiles of conditions associated with increased risk of RSV severe outcomes, stratifying among adults aged 18–59 and ≥60 years.

**Results:** We analyzed data from 1111 adults hospitalized with RSV (median age [IQR] = 66 [53–75]). Among 397 adults aged 18–59 years, two profiles were identified: (1) minimal prevalence with fewer underlying conditions and a posterior median ICU admission risk of 21% (95% credible interval = [16‒25]); (2) cardiorenal/diabetes with frequent heart failure, chronic kidney disease, diabetes, and increased ICU admission risk (37% [27‒48]). Among 714 adults aged ≥60 years, four profiles were identified: (1) minimal prevalence (ICU admission risk = 22% [18‒ 26]), (2) cardiorenal/diabetes (27% [21‒34]), (3) hematologic malignancy and transplant receipt (12% [6‒21]), and (4) chronic pulmonary disease with home oxygen dependence (44% [25‒66]).

**Conclusion:** Distinct underlying condition profiles with varying risks of critical illness were observed among inpatients with RSV. These findings could support recognition of high-risk patients to inform RSV prevention strategies and suggest the role of multimorbidity in severe RSV disease risk warrants further attention.

**Summary:** In this prospective, multicenter analysis of >1,100 adults hospitalized with RSV, patients with multiple underlying conditions were common and partitioned into distinct multimorbidity profiles with varying risks of RSV critical illness.

## Introduction

Respiratory syncytial virus (RSV) is associated with an estimated 123,000–193,000 hospitalizations and 5,000–9,000 deaths annually in US adults, with almost half of all hospitalizations occurring in adults aged ≥75 years [1,2]. Illness severity among adults hospitalized with RSV is similar to severity among hospitalized patients with COVID-19 or influenza [3–6]. Risk factors for RSV hospitalization include older age and underlying conditions (e.g., heart failure, chronic obstructive pulmonary disease [COPD], chronic kidney disease [CKD], severe obesity, and diabetes) [6–13], but there are few data on risk factors for in-hospital outcomes. Additionally, adult patients hospitalized with RSV often present with multiple underlying medical conditions [7,14–16], and few studies have assessed the combined effect of multiple conditions on in-hospital outcome risk.

RSV vaccines were first approved and recommended for older adults in the US in 2023. In 2024, RSV vaccination guidelines were updated to recommend a single vaccine dose for all adults aged ≥75 years and adults aged 60–74 years with medical conditions that confer increased risk of severe disease [1,17,18], and in 2025 recommendations were made for adults aged 50–59 years at increased risk [19]. Characterizing combinations of underlying conditions associated with increased risk of severe RSV disease in adults could inform prevention strategy recommendations [1,17]. Here, we use Bayesian profile regression, a statistical clustering model, to identify profiles of underlying conditions associated with increased risk of severe outcomes among adults hospitalized with RSV during January 2022 through July 2024.

## Methods

### Design and data collection

The Investigating Respiratory Viruses in the Acutely Ill (IVY) network is a multicenter inpatient surveillance network comprising 26 hospitals in 20 U.S. states used for assessing vaccine effectiveness and clinical epidemiology [3,20–23] (Supplementary Materials). During January 2022–July 2024, sites prospectively enrolled inpatients aged ≥18 years who met an acute respiratory illness case definition and had clinical testing performed for SARS-CoV-2, influenza viruses, or RSV (Supplementary Methods). Nasal swab surveillance specimens were additionally collected from all enrolled patients and tested at Vanderbilt University Medical Center for these viruses. Medical records were reviewed by trained personnel who abstracted demographic and clinical data, including underlying conditions and in-hospital outcomes. Receipt of RSV vaccine was identified by electronic medical records, state or city vaccine registries, and plausible self-reported data.

Enrolled inpatients were included if they had RSV infection confirmed by clinical or surveillance testing of a specimen collected within 10 days of symptom onset and 3 days of admission. Patients were excluded if they had co-detection of influenza viruses or SARS-CoV-2 (n = 158), missing underlying conditions (n = 64), or missing outcomes (n = 35). We stratified the cohort by patient age group (18–59 and ≥60 years) to account for age-dependent risk and prevalence of conditions.

### Underlying conditions and severe in-hospital outcomes

We included underlying condition risk factors for severe RSV disease among older adults defined by the Advisory Committee on Immunization Practices [18]. To restrict to the most informative conditions, we removed conditions with high tetrachoric correlation (≥0.7) [24] with other conditions or rare (≤1%) prevalence, resulting in seventeen conditions (Supplementary Methods). Clinical severity was characterized using severe outcomes from presentation to the first of hospital discharge, patient death, or hospital day 28 [3]. These outcomes included (1) intensive care unit (ICU) admission, (2) acute organ failure, defined as either respiratory failure (new receipt of high-flow nasal canula, noninvasive ventilation, or invasive mechanical ventilation [IMV]), cardiovascular failure (vasopressor use), or kidney failure (new receipt of kidney replacement therapy), and (3) a composite of IMV or death (Supplementary Methods).

### Descriptive statistical analyses

We evaluated overall differences using Fisher’s exact test for categorical variables and the Mann-Whitney test for continuous variables. For assessing counts of conditions, conditions were dichotomized, and only severe obesity was included among all body mass index (BMI) categories.

### Bayesian profile regression

We applied Bayesian profile regression, a statistical clustering method that comprises an assignment submodel grouping individuals with a condition profile and a disease submodel that evaluates association of profiles with the outcome. Submodels are jointly fitted in a Bayesian paradigm, enabling the outcome to influence profile membership (Supplementary Methods) [25–28]. We conducted Bayesian profile regression in R (version 4.4.1) separately for the two age groups and three outcome variables using the PReMiuM package [26]. We assessed dependency on initial random profile allocations and outcome variable used (Supplementary Methods). Condition prevalence and severe outcome risk were described using median posterior probabilities and 95% Bayesian equal-tailed credible intervals (i.e., spanning from the 2.5^th^ to 97.5^th^ percentile) [29]. We characterized profiles based on conditions that differed in prevalence among patients within a profile compared to all patients in the age group, as indicated when profile condition prevalence credible intervals excluded empirical prevalence.

Differences in outcome risks relative to the “minimal prevalence” reference profile (defined in Results) were assessed using risk ratios (RR_Profile_); credible intervals that excluded 1 were interpreted as evidence under the Bayesian framework for a difference in outcome risk. We also calculated the posterior probability P(RR_Profile_ > 1).

### Poisson regression

We compared Bayesian profile regression results to frequentist Poisson regression estimates of adjusted risk ratios for individual conditions (RR_Poisson_). To improve estimation for rare outcomes and conditions, we used Firth bias-reduced Poisson regression with robust error variances and the Morel correction as implemented in the *firthb* R package (Supplementary Methods) [30,31]. Models were fit separately for the two age groups and three outcome variables. After applying the Benjamini–Hochberg procedure for multiple testing, two-sided adjusted *P-*values ≤ .05 were interpreted as evidence under a frequentist framework that a condition was significantly associated with a change in outcome risk.

### Ethical review

This activity was reviewed by CDC, deemed not research, and was conducted consistent with applicable federal law and CDC policy. (45 C.F.R. part 46.102(l)(2), 21 C.F.R. part 56; 42 U.S.C. §241(d); 5 U.S.C. §552a; 44 U.S.C. §3501 et seq).

## Results

A total of 1111 RSV inpatients were included from 26 hospitals in 20 U.S. states, comprising 397 adults aged 18–59 and 714 adults aged ≥60. RSV inpatients had a median age of 66 years (IQR 54–75); the majority were female (n = 600; 54.0%) and non-Hispanic White (n = 576; 51.8%) (Table 1). Hospitalization dates ranged from January 17, 2022, through July 5, 2024, and most patients (57.5%) were enrolled during the September 21, 2023‒July 5, 2024 enrollment period. Patients admitted to the ICU had earlier hospital admission relative to symptom onset (median days = 2 [IQR 0–3]) compared to patients not admitted to the ICU (3 [IQR 1–5]; Mann-Whitney *P* <0.001). Ten adults aged ≥60 years (1.4% of adults aged ≥60 years) received RSV vaccination, and 46 patients (4.8% among 968 patients with available data) received ribavirin antiviral treatment during hospitalization. Among 846 patients with known RSV subtype, 373 (44.1%) had subtype A and 473 (55.9%) had subtype B. Underlying conditions were prevalent, with 303 patients (27.3%) having one condition, 274 (24.7%) having two, 224 (20.2%) having three, and 194 (17.5%) having four or more with increasing burden by age (Supplementary Figure 1, Supplementary Tables 1 and 4).

**Table 1.**
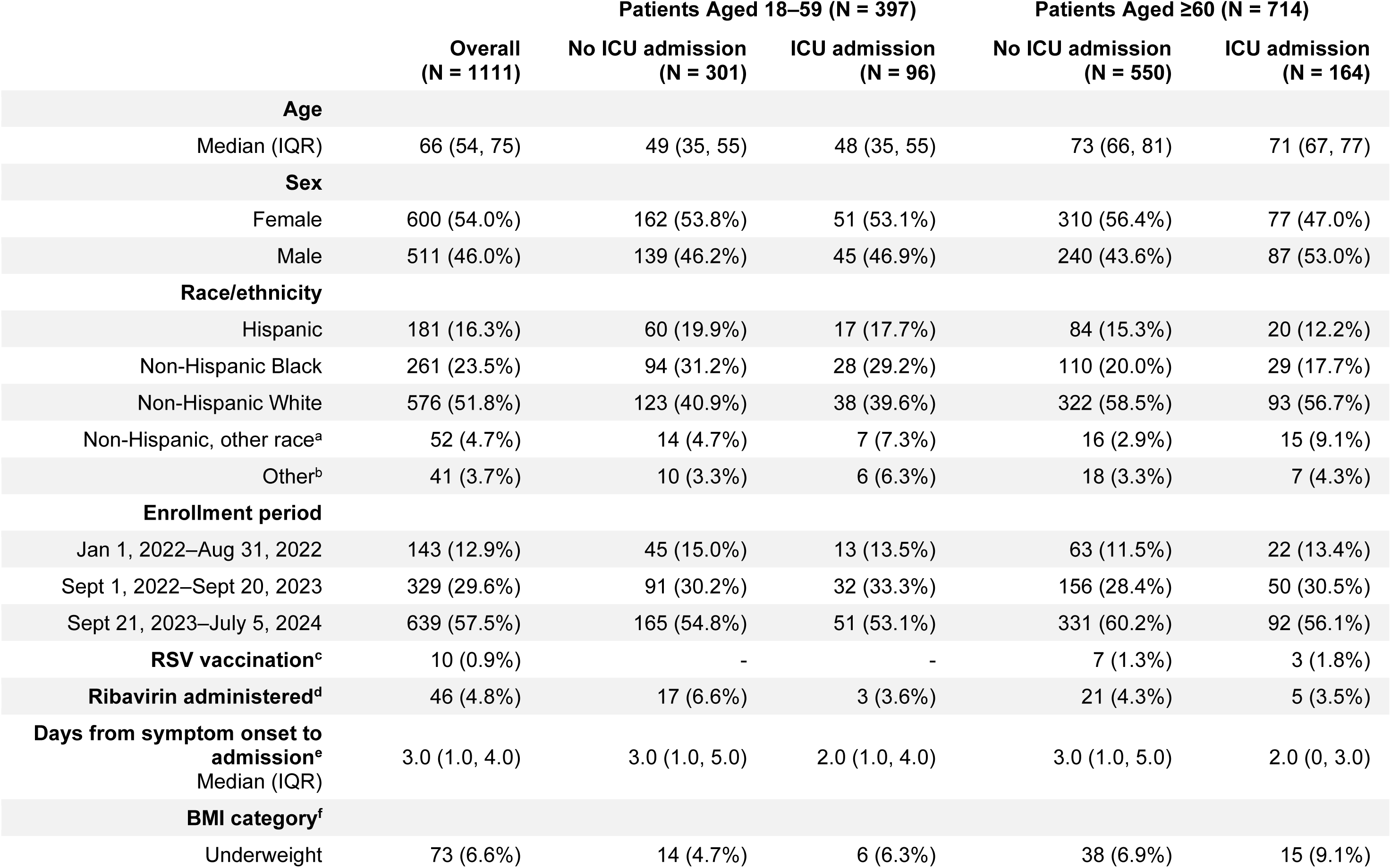
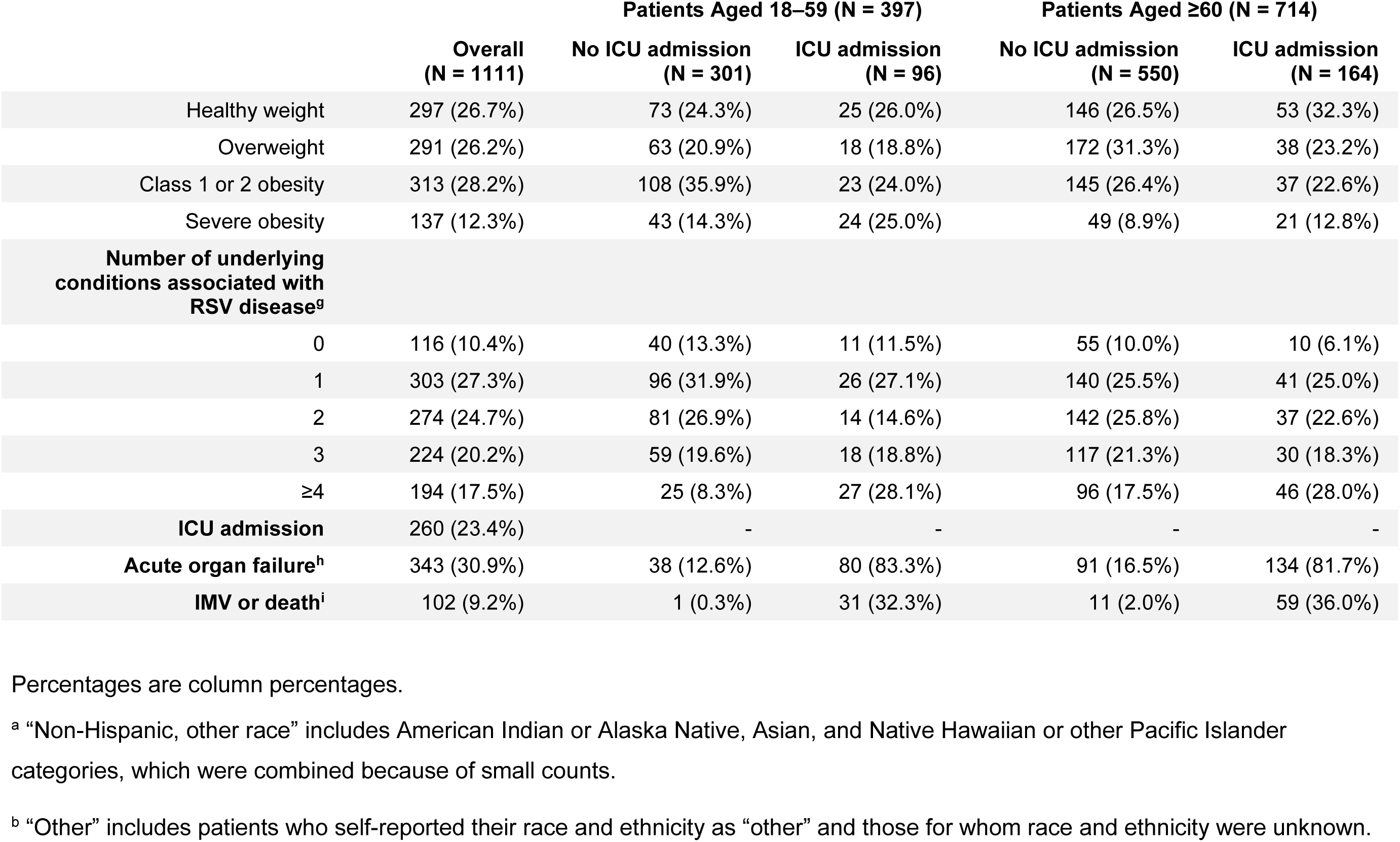
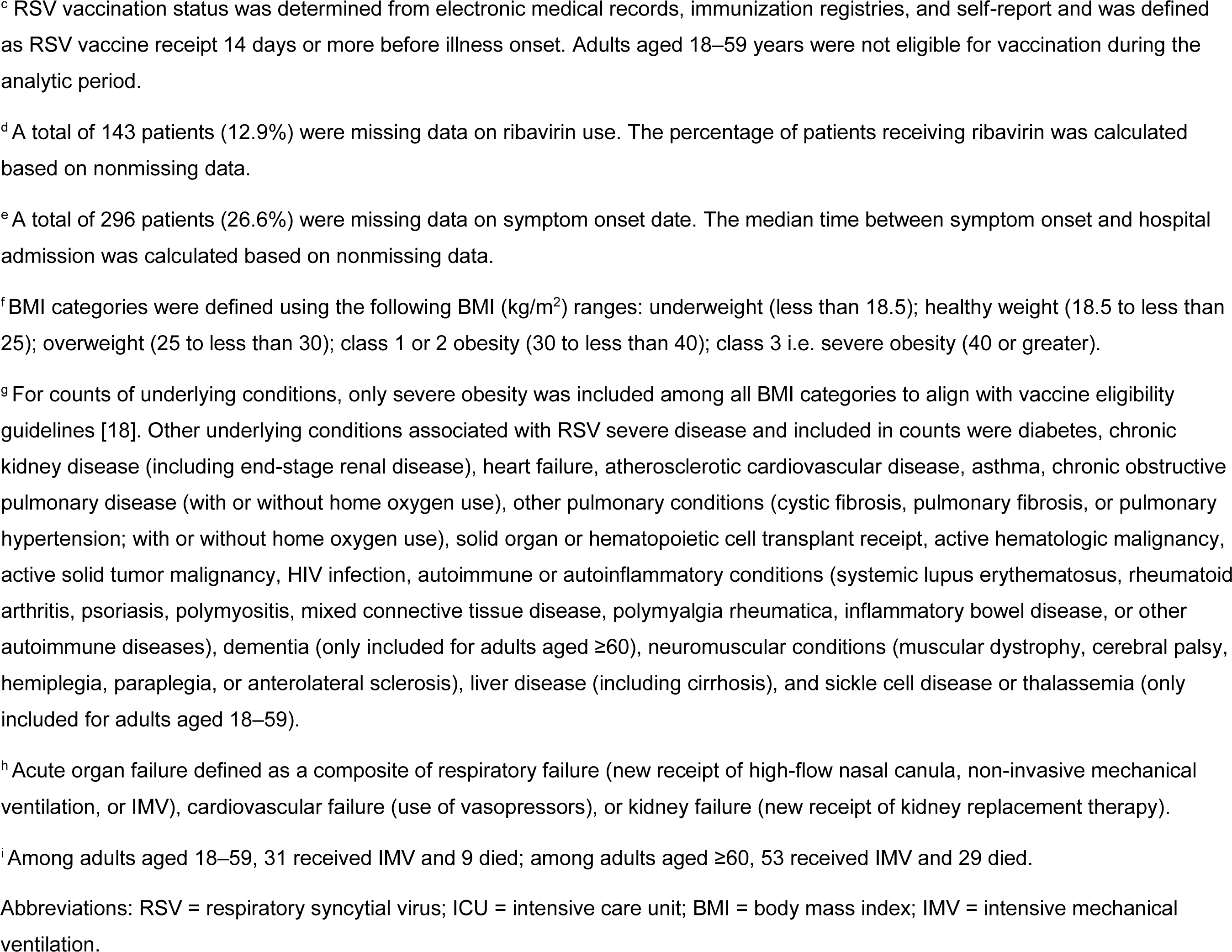
Characteristics of Patients Hospitalized with Respiratory Syncytial Virus by Age Group and Intensive Care Unit Admission Status — IVY Network, 26 Hospitals, January 2022–July 2024, N = 1111.

### Adults aged 18-59 years

Among 397 RSV patients aged 18–59 years (median age = 49 years [IQR 35–55]), 96 (24.2%) were admitted to the ICU, 118 (29.7%) experienced acute organ failure, and 32 (8.1%) received IMV or died (Table 1). Underlying conditions were reported for 346 (87.2%) patients, with the most common being diabetes (n = 109; 27.5%), asthma (n = 96; 24.2%), CKD (n = 87; 21.9%), and heart failure (n = 83; 20.9%) (Figure 1). Patients admitted to the ICU more frequently had severe obesity (25.0% vs. 14.3%; *P =* 0.003) and four or more underlying conditions (28.1% vs. 8.3%; Fisher’s *P =* 0.003). The conditions with the highest positive correlations were heart failure and CKD (tetrachoric correlation = 0.51), CKD and transplant receipt (0.46), and hematologic malignancy and transplant receipt (0.44) (Supplementary Figure 2).

**Figure 1.**
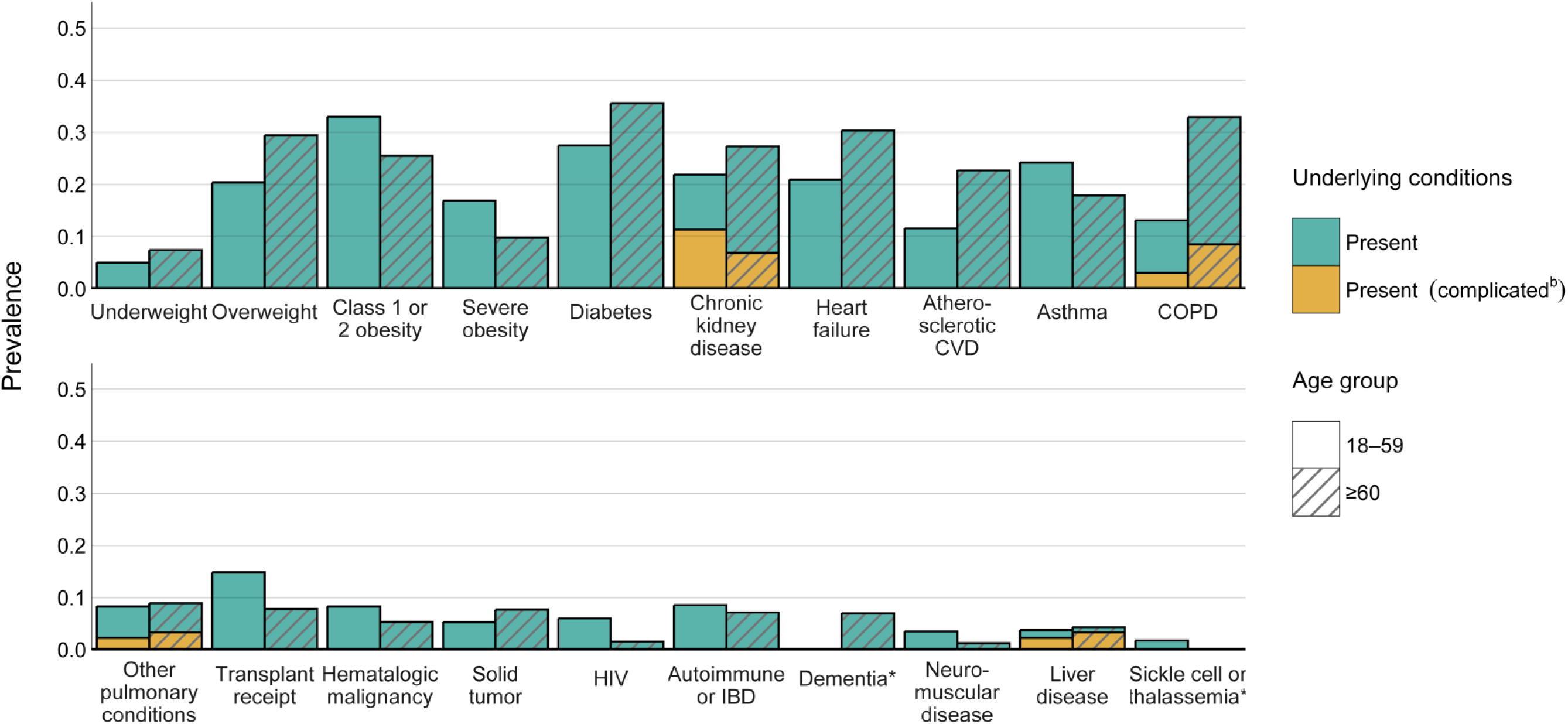
Prevalence of Underlying Conditions^a^ by Age Group Among Patients Hospitalized with Respiratory Syncytial Virus — IVY Network, 26 Hospitals, January 2022–July 2024, N = 1111 * The prevalence of dementia in adults aged 18‒59 and sickle cell or thalassemia in adults aged ≥60 was <1%. ^a^ BMI categories were defined using the following BMI (kg/m^2^) ranges: underweight (less than 18.5); healthy weight (18.5 to less than 25); overweight (25 to less than 30); class 1 or 2 obesity (30 to less than 40); class 3 i.e. severe obesity (40 or greater). Underlying conditions potentially associated with RSV disease included underweight, overweight, class 1 or 2 obesity, severe obesity, diabetes, chronic kidney disease (including end-stage renal disease), heart failure, atherosclerotic cardiovascular disease, asthma, chronic obstructive pulmonary disease (with or without home oxygen use), other pulmonary conditions (cystic fibrosis, pulmonary fibrosis, or pulmonary hypertension; with or without home oxygen use), solid organ or hematopoietic cell transplant receipt, active hematologic malignancy, active solid tumor malignancy, HIV infection, autoimmune or autoinflammatory conditions (systemic lupus erythematosus, rheumatoid arthritis, psoriasis, polymyositis, mixed connective tissue disease, polymyalgia rheumatica, inflammatory bowel disease, or other autoimmune diseases), dementia (only included for adults aged ≥60), neuromuscular conditions (muscular dystrophy, cerebral palsy, hemiplegia, paraplegia, or anterolateral sclerosis), liver disease (including cirrhosis), and sickle cell disease or thalassemia (only included for adults aged 18‒59). ^b^ Complicated conditions include COPD with home oxygen use, other pulmonary conditions with home oxygen use, liver disease with cirrhosis, and chronic kidney disease with end stage renal disease as indicated by chronic renal replacement therapy. Abbreviations: ICU = intensive care unit; IMV = intensive mechanical ventilation; BMI = body mass index; CVD = cardiovascular disease; COPD = chronic obstructive pulmonary disease; IBD = inflammatory bowel disease.

Using Bayesian profile regression, we identified two patient profiles. Patients with the first profile constituted 78% (n = 308) of the cohort; we termed this the *minimal prevalence* profile as patients had fewer conditions than overall patients in this age group (median number = 1 [IQR 1–2]; Figure 2A, Supplementary Tables 1 and 2). Patients with the second profile constituted the remaining 22% (n = 89) of the cohort and had increased multimorbidity (median number of conditions = 3 [IQR 3–4]). Prevalences of heart failure (posterior median [95% credible interval] = 47.7% [36.9‒58.7]), diabetes (41.3% [31.8‒51.1]), and CKD with (29.6% [19.9‒40.5]) and without (22.9% [14.9‒32.3]) end-stage renal disease were increased relative to empiric prevalences. We termed this the *cardiorenal and diabetes* profile as the majority (71%) of patients had two or more of these three conditions (Supplementary Table 3).

**Figure 2.**
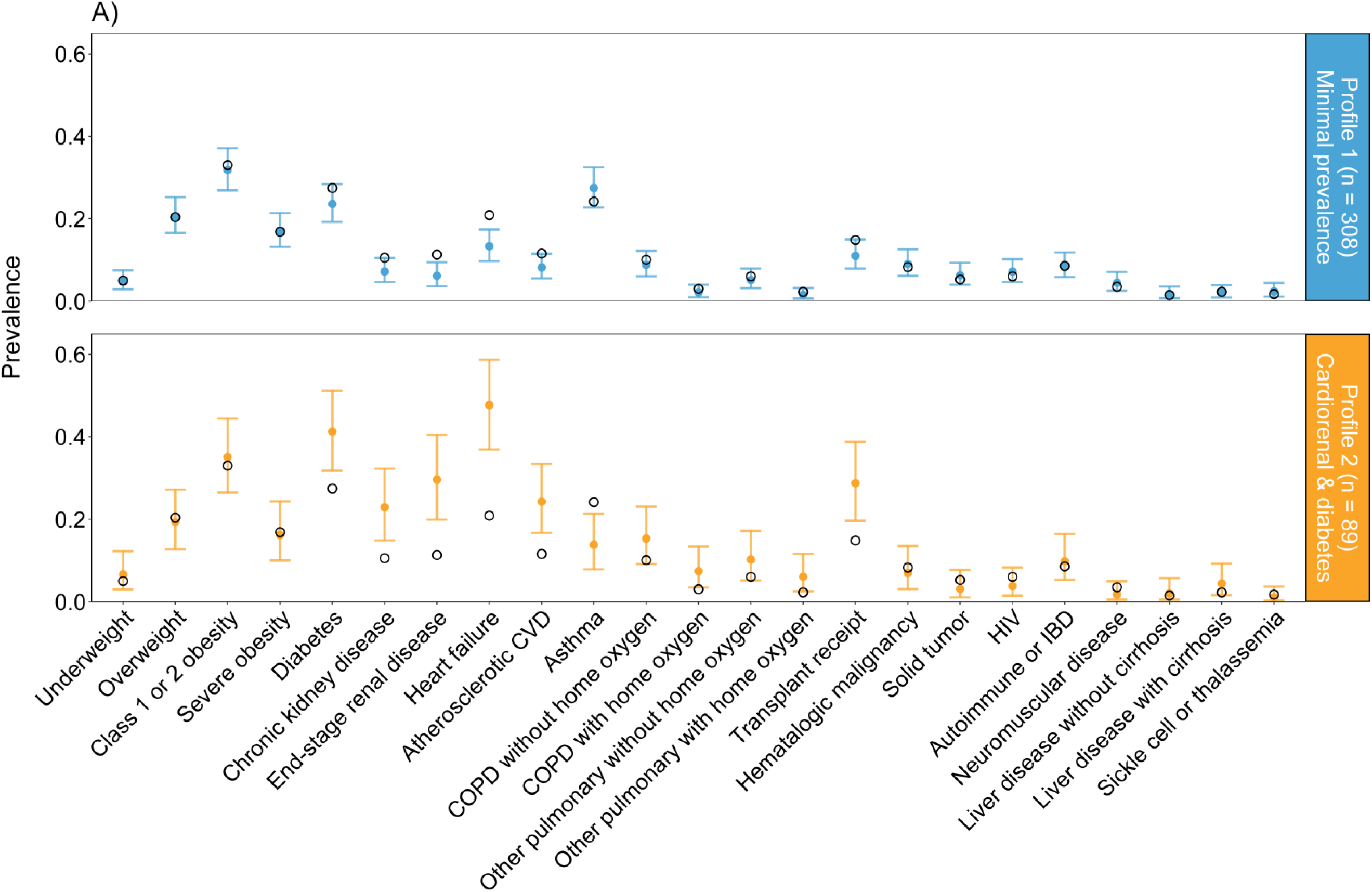
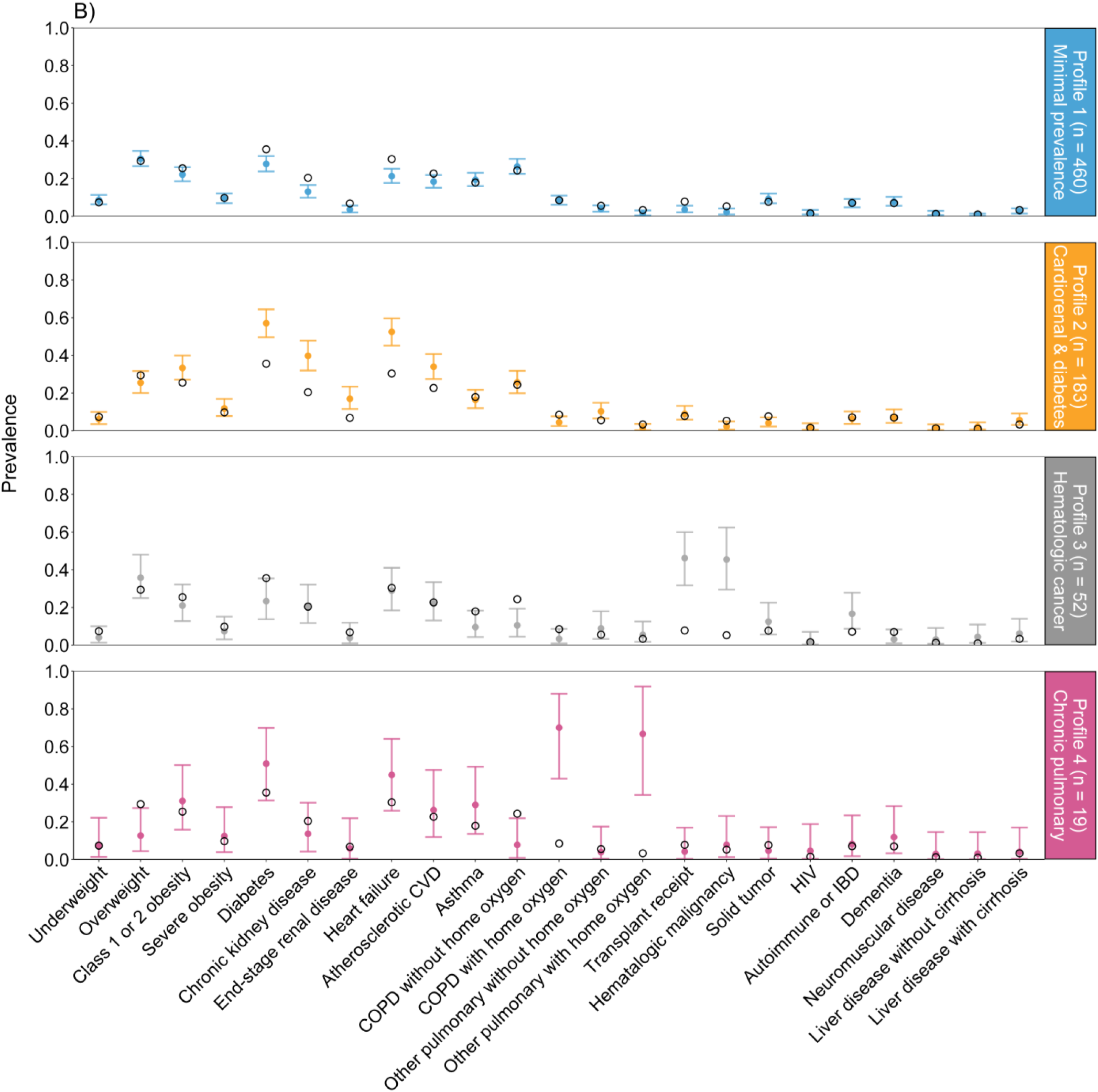
Prevalence and 95% Credible Intervals of Underlying Conditions^a^ By Bayesian Profiles^b^ of Patients A) Aged 18–59 (N = 397) and B) ≥60 (N = 714) Hospitalized with Respiratory Syncytial Virus — IVY Network, 26 Hospitals, January 2022– July 2024 Closed circles and error bars are median prevalence estimates and 95% Bayesian equal-tailed credible intervals from the posterior distribution, respectively. Open circles indicate empirical prevalence of the condition among all patients within the age group. Among adults in both age groups, profile 1 represents the minimal prevalence profile and 2 represents the cardiorenal and diabetes profile. Among adults aged ≥60, profile 3 represents the hematologic malignancy profile, and 4 represents the severe chronic pulmonary disease with home oxygen dependence profile. ^a^ BMI categories were defined using the following BMI (kg/m^2^) ranges: underweight (less than 18.5); healthy weight (18.5 to less than 25); overweight (25 to less than 30); class 1 or 2 obesity (30 to less than 40); class 3 i.e. severe obesity (40 or greater). Underlying conditions potentially associated with RSV disease included underweight, overweight, class 1 or 2 obesity, severe obesity, diabetes, chronic kidney disease (including end-stage renal disease), heart failure, atherosclerotic cardiovascular disease, asthma, chronic obstructive pulmonary disease (with or without home oxygen use), other pulmonary conditions (cystic fibrosis, pulmonary fibrosis, or pulmonary hypertension; with or without home oxygen use), solid organ or hematopoietic cell transplant receipt, active hematologic malignancy, active solid tumor malignancy, HIV infection, autoimmune or autoinflammatory conditions (systemic lupus erythematosus, rheumatoid arthritis, psoriasis, polymyositis, mixed connective tissue disease, polymyalgia rheumatica, inflammatory bowel disease, or other autoimmune diseases), dementia (only included for adults aged ≥60), neuromuscular conditions (muscular dystrophy, cerebral palsy, hemiplegia, paraplegia, or anterolateral sclerosis), liver disease (including cirrhosis), and sickle cell disease or thalassemia (only included for adults aged 18‒59). ^b^ Bayesian profile regression is an outcome variable-informed statistical clustering approach that comprises an assignment submodel (assigns individuals to a condition profile) and a disease submodel (evaluates association of profiles with the outcome variable); the two submodels are jointly fitted in a Bayesian paradigm, enabling the outcome variable to influence profile membership. Abbreviations: ICU = intensive care unit; IMV = intensive mechanical ventilation; BMI = body mass index; CVD = cardiovascular disease; COPD = chronic obstructive pulmonary disease; IBD = inflammatory bowel disease.

Patients with the cardiorenal and diabetes profile had elevated risk of ICU admission (37.1% [27.3‒47.6]; relative risk RR_Profile_ = 1.81 [1.26‒2.56]) compared to patients with the minimal prevalence profile (20.5% [16.1‒25.0]) (Figure 3). Relative risks of acute organ failure (37.2% [28.1‒46.4]; RR_Profile_ = 1.37 [1.00‒1.82]) and IMV or death (10.9% [5.7‒17.7]; RR_Profile_ = 1.51 [0.70‒2.88]) were also elevated, though credible intervals for IMV or death did not exclude the null (Supplementary Table 4). Patients with the minimal prevalence profile tended to be slightly younger (median age = 48 years [IQR 34–54]) compared to patients with the cardiorenal and diabetes profile (51 years [IQR 46–56]; Mann-Whitney *P* <0.001) (Supplementary Figure 3).

**Figure 3.**
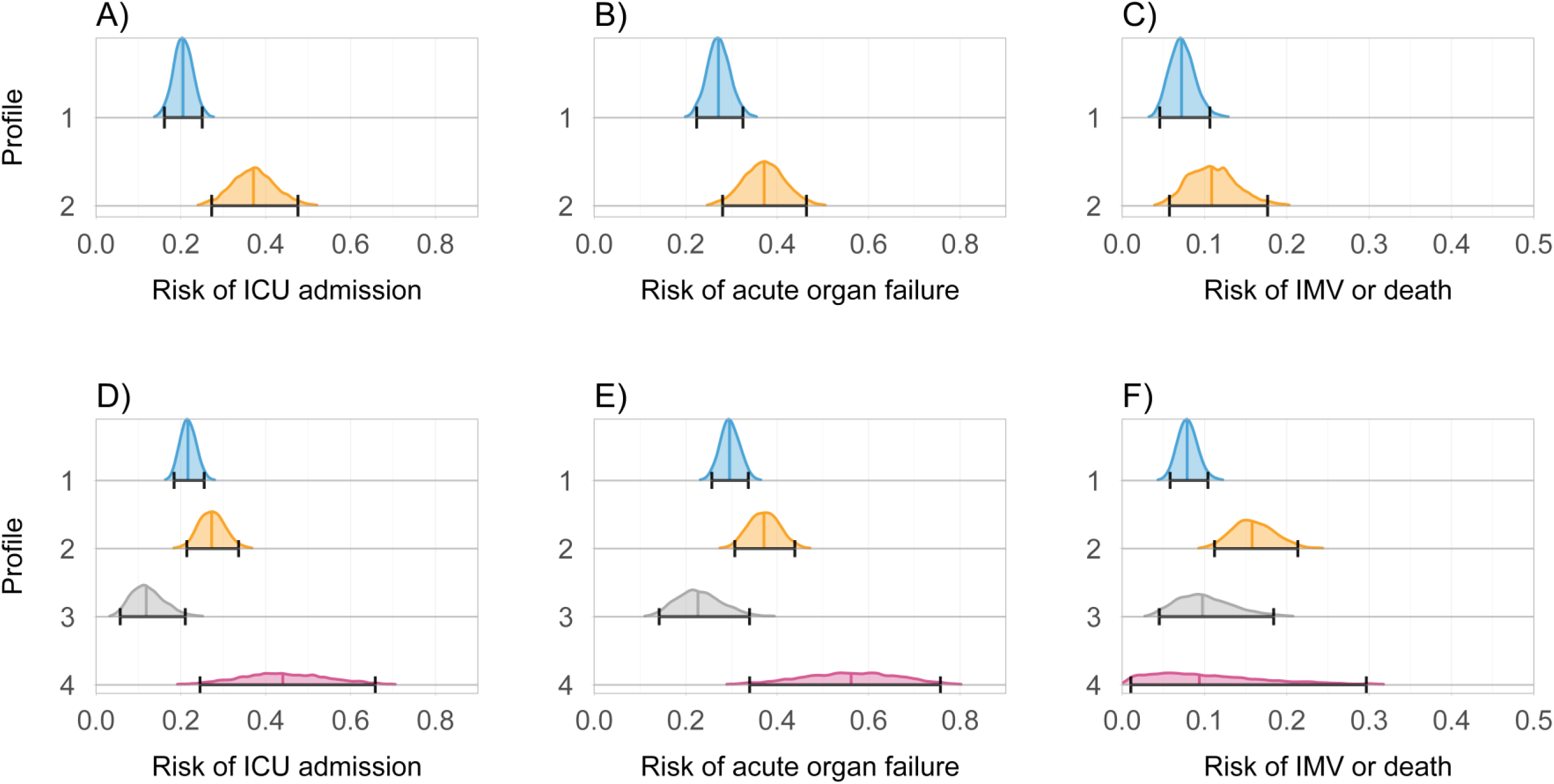
Risk of Severe Outcomes^a^ and 95% Credible Intervals Among Bayesian Profiles^b^ of Patients Hospitalized with Respiratory Syncytial Virus Aged 18–59 (N = 397; A–C) and ≥60 (N = 714; D–F) Years — IVY Network, 26 Hospitals, January 2022–July 2024 Posterior distributions are depicted by equal-area density plots with taller regions indicating greater probability density. Median posterior distribution estimates are represented by the colored vertical line and error bars are 95% Bayesian equal-tailed credible intervals. Among adults in both age groups, profile 1 represents the minimal prevalence profile and 2 represents the cardiorenal and diabetes profile. Among adults aged ≥60, profile 3 represents the hematologic malignancy profile, and 4 represents the severe chronic pulmonary disease with home oxygen dependence profile. ^a^ Severe outcomes included ICU admission, acute organ failure, and invasive mechanical ventilation (IMV) or death. Acute organ failure was defined as a composite of respiratory failure (new receipt of high-flow nasal canula, non-invasive mechanical ventilation, or IMV), cardiovascular failure (use of vasopressors), or kidney failure (new receipt of kidney replacement therapy). ^b^ Bayesian profile regression is an outcome variable-informed statistical clustering approach that comprises an assignment submodel (assigns individuals to a condition profile) and a disease submodel (evaluates association of profiles with the outcome variable); the two submodels are jointly fitted in a Bayesian paradigm, enabling the outcome variable to influence profile membership. Abbreviations: ICU = intensive care unit; IMV = intensive mechanical ventilation.

### Adults aged ≥60 years

Among 714 RSV patients aged ≥60 (median age = 72 years [IQR 66–80]), 164 patients (23.0%) were admitted to the ICU, 225 (31.5%) experienced acute organ failure, and 70 (9.8%) received IMV or died. Underlying conditions were reported for 649 (90.9%) patients; the most common were diabetes (n = 254; 35.6%), COPD (n = 235; 32.9%), heart failure (n = 217; 30.4%), and CKD (n = 195; 27.3%) (Figure 1). Patients admitted to the ICU more frequently had severe obesity (12.8% vs 8.9%; Fisher’s *P <*0.001) and four or more underlying conditions (28.0% vs. 17.5%; Fisher’s *P =* 0.004) (Table 1). The most positively correlated conditions were hematologic malignancy and transplant receipt (0.699), CKD and heart failure (0.44), and CKD and diabetes (0.38) (Supplementary Figure 2).

We identified four profiles using Bayesian profile regression. Profile one (*minimal prevalence*, as for younger adults) comprised 460 (64%) patients with fewer conditions (median number = 1 [IQR 1–2]; Supplementary Table 5 and 6, Figure 2B). Profile two (*cardiorenal and diabetes*) comprised 183 patients (26%) with increased multimorbidity (median number of conditions = 4 [IQR 3–4]) and higher prevalence of diabetes (57.1% [49.6‒64.4]), heart failure (52.5% [45.2‒59.7]), and CKD with (17.0% [11.5‒23.4]) and without (39.8% [32.0‒47.8]) end-stage renal disease; 87% of patients in this profile had two or more of these conditions (Supplementary Table 3). Profile three comprised 52 patients (7%; median number of conditions = 3 [IQR 2–4]) with higher prevalence of hematologic malignancy (45.4% [29.5‒62.4]) and transplant receipt (46.2% [31.7‒60.0]); we termed this the *hematologic malignancy* profile. Profile four comprised the remaining 19 patients (3%; median number of conditions = 4 [IQR 3–5]) who had increased prevalence of both COPD (70.0% [42.9‒88.0]) and other pulmonary conditions (n = 15 with pulmonary hypertension and n = 4 with pulmonary fibrosis) with home oxygen dependence (66.7% [34.3‒91.8]), indicative of severe chronic pulmonary disease.

Patients with the severe chronic pulmonary disease profile experienced the highest ICU admission risk (posterior median [95% credible interval] = 44.0% [24.5‒65.8]), followed by the cardiorenal and diabetes (27.3% [21.4‒33.6]), minimal prevalence (21.6% [18.4‒25.5]), and hematologic malignancy profiles (11.8% [5.7‒21.0]) (Figure 3). Relative to the minimal prevalence profile, patients with the severe chronic pulmonary disease profile had a higher risk of ICU admission (RR_Profile_ = 2.03 [1.11‒3.12]) and acute organ failure (1.90 [1.15‒2.65]) (Supplementary Table 3). Patients with the cardiorenal and diabetes profile had a higher risk of ICU admission (1.26 [0.96‒1.62]), acute organ failure (1.25 [0.99‒1.56]), and IMV or death (2.00 [1.31‒3.04]); credible intervals included the null for the first two outcomes. Patients with the hematologic malignancy profile had a decreased risk of ICU admission (0.55 [0.26‒0.98]). Compared to all other patients, patients in the hematologic malignancy group more frequently received ribavirin antiviral treatment (37.2% vs. 1.7% among patients with available data; Fisher’s *P* <0.001) (Supplementary Table 5).

### Individual risk conditions

Using Poisson regression for evaluating individual conditions, among adults aged 18–59 years the most significant association was COPD without home oxygen use and acute organ failure (RR_Poisson_ = 2.14, 95% confidence interval = [1.52, 3.00]; Benjamini-Hochberg adjusted *P-*value <0.001) (Figure 4, Supplementary Figure 4). This association persisted stratifying within Bayesian inpatient profiles, and the highest organ failure risk was observed among patients with the cardiorenal and diabetes profile and COPD (Supplementary Table 7). Among adults aged ≥60 years, the most significant association was COPD with home oxygen dependence and acute organ failure (RR_Poisson_ = 2.21 [1.60, 3.07]; Benjamini-Hochberg adjusted *P-*value <0.001) (Figure 4, Supplementary Figure 5).

**Figure 4.**
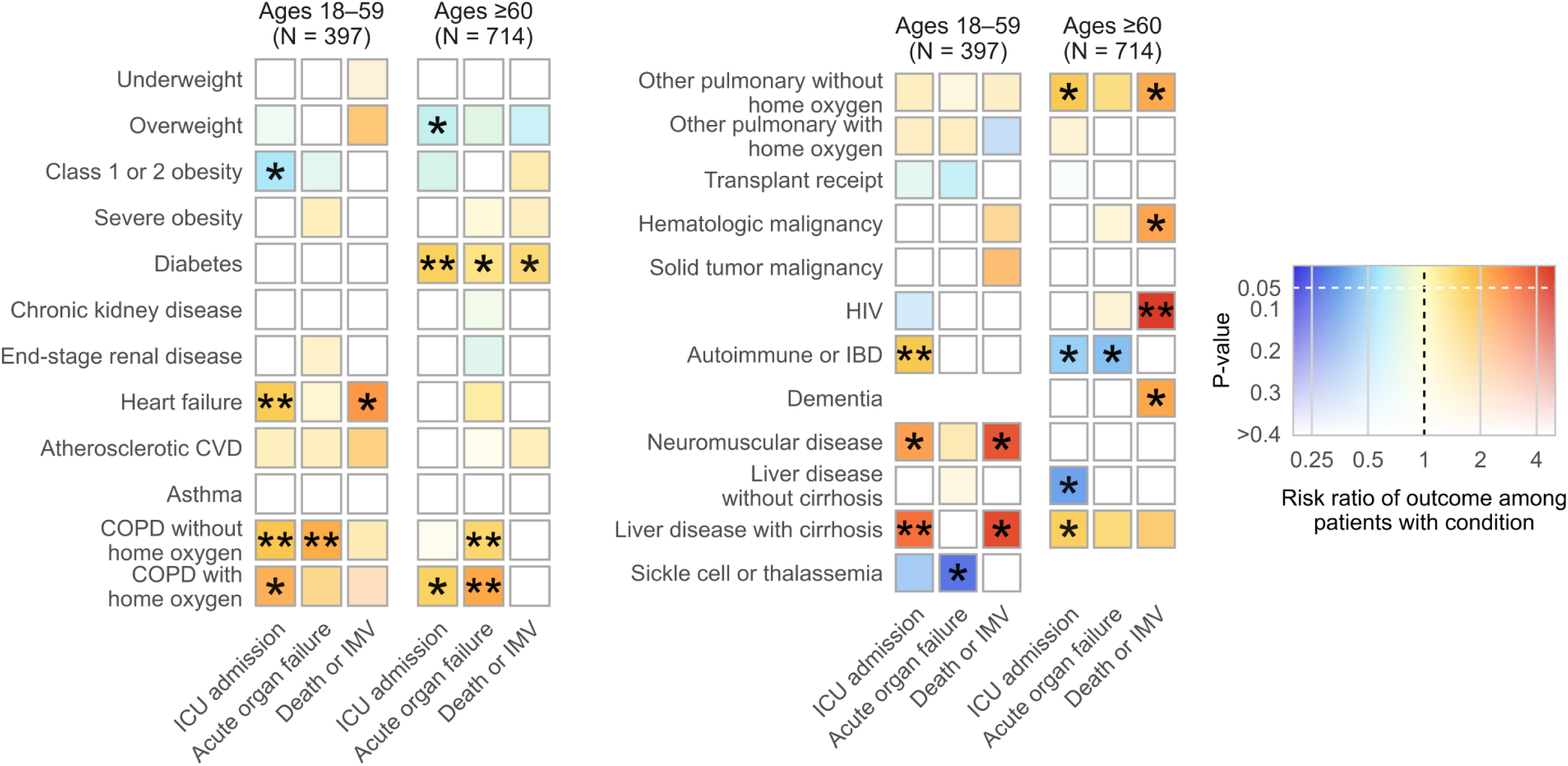
Heatmap of *P-*value (Opacity) Versus Risk^a^ (Hue) of Severe In-Hospital Outcomes^b^ by Underlying Condition^c^ Among Patients Hospitalized with Respiratory Syncytial Virus Estimated Using Poisson Regression — IVY Network, 26 Hospitals, January 2022–July 2024 * indicates two-sided *P-*values ≤ .05 unadjusted for multiple testing ** indicates two-sided *P-*values ≤ 0.05 adjusted for multiple testing correction using the Benjamini-Hochberg procedure with a false-discovery rate of 0.05 ^a^ Risk ratios were estimated using Firth bias-reduced Poisson regression models with robust error variances [30] separately for the two age groups and three outcome variables. Hue indicates the sign and magnitude of the point estimate of the risk ratio (blues for decreased risk, reds for increased risk) and opacity indicates *P-*value (increasing opacity for decreasing *P-*values). ^b^ Severe outcomes included ICU admission, acute organ failure, and invasive mechanical ventilation (IMV) or death. Acute organ failure was defined as a composite of respiratory failure (new receipt of high-flow nasal canula, non-invasive mechanical ventilation, or IMV), cardiovascular failure (use of vasopressors), or kidney failure (new receipt of kidney replacement therapy). ^c^ BMI categories were defined using the following BMI (kg/m^2^) ranges: underweight (less than 18.5); healthy weight (18.5 to less than 25); overweight (25 to less than 30); class 1 or 2 obesity (30 to less than 40); class 3 i.e. severe obesity (40 or greater). Underlying conditions potentially associated with RSV disease included underweight, overweight, class 1 or 2 obesity, severe obesity, diabetes, chronic kidney disease (including end-stage renal disease), heart failure, atherosclerotic cardiovascular disease, asthma, chronic obstructive pulmonary disease (with or without home oxygen use), other pulmonary conditions (cystic fibrosis, pulmonary fibrosis, or pulmonary hypertension; with or without home oxygen use), solid organ or hematopoietic cell transplant receipt, active hematologic malignancy, active solid tumor malignancy, HIV infection, autoimmune or autoinflammatory conditions (systemic lupus erythematosus, rheumatoid arthritis, psoriasis, polymyositis, mixed connective tissue disease, polymyalgia rheumatica, inflammatory bowel disease, or other autoimmune diseases), dementia (only included for adults aged ≥60), neuromuscular conditions (muscular dystrophy, cerebral palsy, hemiplegia, paraplegia, or anterolateral sclerosis), liver disease (including cirrhosis), and sickle cell disease or thalassemia (only included for adults aged 18‒59). Abbreviations: ICU = intensive care unit; IMV = intensive mechanical ventilation; BMI = body mass index; CVD = cardiovascular disease; COPD = chronic obstructive pulmonary disease; IBD = inflammatory bowel disease.

## Discussion

In this prospective, multicenter analysis of >1,100 adults hospitalized with RSV, 90% of patients had at least one underlying condition and almost two thirds had multiple. These conditions partitioned into two profiles for adults aged 18–59 years and four for adults ≥60 years. Across both age groups, approximately one in four inpatients with RSV had a profile characterized by elevated heart failure, diabetes, and CKD; patients with this cardiorenal and diabetes profile had increased risk of multiple severe in-hospital outcomes. Among adults aged ≥60 years, a rare severe chronic pulmonary disease with home oxygen dependence profile was identified with the highest risk of ICU admission and acute organ failure. A separate profile of patients with prevalent hematologic cancer or transplant receipt was also observed with lower ICU admission risk. The majority of patients in both age groups partitioned into a profile with few underlying conditions but an ICU admission risk of ∼21%, highlighting how many patients hospitalized for RSV are still at risk for progression to severe outcomes irrespective of age or multimorbidity.

Using Poisson regression for individual risk conditions, we validate COPD as a risk factor for acute organ failure among adults in both age groups [32]. Taken together, these findings identify subgroups of hospitalized adults who may have increased risk of severe RSV disease, which could inform healthcare provider counseling on RSV vaccination for currently eligible adults and future RSV prevention strategies for adults aged <60 years.

Few studies have examined multimorbidity among adults with RSV disease or characterized the role of multimorbidity in increasing risk for severe outcomes. An analysis from a large US population-based network found that adults with more chronic conditions had higher rates of RSV hospitalization [7]. This and other analyses have primarily categorized multimorbidity by condition count or indices (e.g. Charlson or multimorbidity-weighted) [7,14–16]. Here, we characterized specific concurrent conditions that jointly were associated with increased severity. The triad of heart failure, CKD, and diabetes that we observed is a well-recognized clinical profile among adults [33–36]. In a study of >87,000 adults with newly diagnosed heart failure in the United Kingdom, 16% had concurrent CKD and diabetes and 52% had either condition; patients with CKD, diabetes, or both had higher rates of hospitalizations and death than those with heart failure alone [36]. Prior studies have also identified heart failure, CKD, and diabetes as individual risk factors for RSV-associated hospitalization [8,13,37]. Our study advances these findings by demonstrating the potential of concurrent heart failure, CKD, and diabetes as a combined RSV risk factor.

This analysis used Bayesian profile regression, developed to address limitations of traditional regression methods for modeling correlated covariates [25,38]. We demonstrated how Bayesian and traditional regression methods can provide complementary insights. Among adults ≥60 years, COPD was identified as a primary risk factor using Poisson regression, in line with prior studies [32]. Extending this, profile regression highlighted an infrequent subgroup of COPD patients with home oxygen dependence and other concurrent pulmonary conditions, including pulmonary hypertension, a complication of COPD [39]. COPD was also identified as a risk factor among younger adults, but did not coincide directly with a Bayesian profile. Instead, we observed evidence for additive risk contributions from factors identified independently using Bayesian and traditional regression, with elevated acute organ failure risk occurring among cardiorenal and diabetes profile inpatients who also had COPD. These findings emphasize how risk for severe outcomes can vary across a complex landscape of underlying conditions and support the use of diverse methodologic approaches for risk factor characterization.

The findings in this report are subject to several limitations. First, the profiles generated by Bayesian profile regression are exploratory, subject to misclassification, and require validation. Follow up analyses are warranted to construct more formal clinical subgroup definitions and compare how definitions vary in sensitivity for predicting severe outcomes. Second, our profile labels are simplified descriptors that may not fully capture all features or causal risk factors.

Third, we did not adjust for confounders beyond stratifying by age, and limited sample sizes precluded further stratification. Fourth, this analysis conditions on hospitalization and was not designed to assess risk factors for RSV-associated hospitalization, including age. Thus, although we found that inpatients aged ≥60 years with a hematologic malignancy profile had a lower ICU admission risk than patients with a minimal prevalence profile, this finding does not negate immunocompromising conditions as risk factors for RSV-associated hospitalization [18]. The observed reduced risk of ICU admission may reflect differences in clinical thresholds for hospital admission [13] or ribavirin antiviral treatment practices among patients with hematologic malignancies or hematopoietic cell transplants [40,41], though further investigation is needed.

In summary, distinct profiles of underlying conditions with varying risks of severe outcomes were observed among adults hospitalized with RSV. Adults hospitalized with RSV of all ages with a cardiorenal and diabetes profile and adults aged ≥60 years with chronic pulmonary disease and home oxygen dependence were at increased risk for severe outcomes. Findings from this analysis suggest that the role of multimorbidity in increasing severe RSV disease risk warrants further attention.

## Data Availability

No additional data available.

## Acknowledgments.

Michael Melgar, MD (CDC) and Amadea Britton, MD (CDC) for critical input. Ryan Wiegand, PhD (CDC), Lan Shi, MS (VUMC), Trey McGonigle, MS (VUMC), and Bryan Blette, PhD (VUMC) for statistical review. These individuals were not compensated for their work.

