## Supplementary Materials for "Multimorbidity Profiles and Severe In-Hospital Outcomes in Adults with Respiratory Syncytial Virus"

IVY

Investigators and Collaborators……………………………………………………………………3

Supplementary Methods…………………………………………………………………………6

Supplementary Tables and Figures

- Supplementary Table 1. Characteristics and Number of Underlying Conditions Associated with Respiratory Syncytial Virus Disease Among Bayesian Profiles of 397 Patients Aged 18–59 Hospitalized with Respiratory Syncytial Virus……………………..15
- Supplementary Table 2. Prevalence of Underlying Conditions Associated with Respiratory Syncytial Virus Disease Among Bayesian Profiles of 397 Patients Aged 18–59 Hospitalized with Respiratory Syncytial Virus…………………………………………..18
- Supplementary Table 3. Prevalence of Heart Failure, Chronic Kidney Disease, and Diabetes Among Bayesian Cardiorenal and Diabetes Profile of Patients Hospitalized with Respiratory Syncytial Virus by Age Group……………………………………………21
- Supplementary Table 4. Risk Ratios and Posterior Probabilities of Severe Outcomes Among Bayesian Profiles of Patients Hospitalized with Respiratory Syncytial Virus……………………………………………………………………………………..22
- Supplementary Table 5. Characteristics and Number of Underlying Conditions Associated with Respiratory Syncytial Virus Disease Among Bayesian Profiles of 714 Patients Aged ≥60 Hospitalized with Respiratory Syncytial Virus ……………………….24

- Supplementary Figure 4. Risk of Severe In-Hospital Outcome by Condition Among 397 Patients Aged 18–59 Hospitalized with Respiratory Syncytial Virus Estimated Using Poisson Regression ……………………………………………………………..…………37

- Supplementary Figure 5. Risk of Severe In-Hospital Outcome by Condition Among 714 Patients Aged ≥60 Hospitalized with Respiratory Syncytial Virus Estimated Using Poisson Regression…………………………………………………………………...……39

- Supplementary References……………………………………………………...………...41

**IVY Network Group**

Investigators and collaborators of the Investigating Respiratory Viruses in the Acutely Ill (IVY) Network are listed below.

**Baylor, Scott and White, Temple and Dallas, Texas**
Manju Gaglani, Shekhar Ghamande, Tresa McNeal, Cristie Columbus, Robert L. Gottlieb, Catherine Raver, Ashley Bychkowsky, Symone Dunkley, Tammy Fisher, Daniela Gonzalez, Therissa Grefsrud, Mariana Hurutado-Rodriguez, Gabriela Perez

**Baylor University Medical Center, Dallas, Texas**
Ashley Bychkowski, Symone Dunkley, Tammy Fisher, Daniela Gonzalez, Therissa Grefsrud, Mariana Hurutado-Rodriguez, Gabriela Perez

**Baystate Medical Center, Springfield, Massachusetts**
Jay Steingrub, Lesley De Souza, Scott Ouellette, Cynthia Kardos, Rae Lynn Defeo

**Beth Israel Medical Center, Boston Massachusetts**
Nathan I. Shapiro, Michael Bolstad, Brianna Coviello, Robert Ciottone, Arnaldo Devilla, Ana Grafals, Conor Higgins, Carlo Ottanelli, Kimberly Redman, Douglas Scaffidi, Alexander Weingart

**Centers for Disease Control and Prevention (CDC), Atlanta, Georgia**
Diya Surie, Kevin C. Ma, Fatimah S. Dawood, Nathaniel M. Lewis, Sascha Ellington

**Cleveland Clinic, Cleveland, Ohio**
Omar Mehkri, Megan Mitchell, Zachary Griffith, Connery Brennan, Kiran Ashok, Bryan Poynter, Abhijit Duggal

**Emory University, Atlanta, Georgia**
Laurence Busse, William Bender, Caitlin ten Lohuis

**Hennepin County Medical Center, Minneapolis, Minnesota**
Mary O'Rourke, Leyla Taghizadeh, Laurynn Giles, Audrey Hendrickson, Anne Frosch

**Henry Ford Health, Detroit, Michigan**
Ivana A Vaughn, Mayur Ramesh, Lois E Lamerato, Ishraaq Atkins, Jaleesa Clark, Alycia Lilla, Catherine McKeon, Rachna Jayaprakash, Sindhuja Koneru, Jean Ashley Lava, Zina Pinderi, Melissa Resk, Shruti Tirumala, Katrina Williams

**Intermountain Medical Center, Murray, Utah**
Ithan D. Peltan, Samuel M. Brown, Joslyn Bassett, Shandi Poulson, Vineela Thumma

**Johns Hopkins University, Baltimore, Maryland**
David N. Hager, Harith Ali, Safa Saeed

**Montefiore Medical Center, Bronx, New York**
Michelle Gong, Amira Mohamed

**Ohio State Medical Center, Columbus, Ohio**
Maryiam Khan, Gabrielle Swoope, Sarah Karow, Brooke Lee, Amer Charif, Ibrahim Abu Hammad, Connor Lang, Kristina Luikart, Amanie Rasul, Rasha Alrifae, Rashil Madan, Jun Park, Madison So, Preston So, David Smith, Connor Snyder, Reece Wilson, Zachery Lewald, Manisha Pathak, Elli Schwartz

**Oregon Health and Sciences University, Portland, Oregon**
Adrian Hernandez-Frausto, Edvinas Pocius, Emily Tribbett, Genesis Briceno, Jose Pena, Alex Vazquez-Cortes, Anna Romo, Annabelle Blue, Connie Tran, Faith Crane, Jenny Chan, Maria Makman, Amy Segura, Taylor Nichols, Yamilenia Aguilar Diaz

**Stanford University, Stanford, California**
Cynthia Perez, Grace Kyin-Ye Tam, Lily Lau, Vanessa Pitre, Samantha Ferguson, Jennifer G. Wilson, Leonard Basobas, Alexandra Gordon

**University of Arizona, Tucson, Arizona**
Cameron Hypes, Beth Salvagio Campbell, Karen Lutrick

**University of California-Los Angeles, Los Angeles, California**
Cody Tran, Sukantha Chandrasekaran, Omai Garner

**University of Colorado, Aurora, Colorado**
Adit A. Ginde, Samantha Simon, Amanda Martinez, Amy Sullivan, Laura Aguilar-Marquez, Erika Alor, Yvette Evans, Jacob Rademacher

**University of Iowa, Iowa City, Iowa**
Nicholas Mohr, Anne Zepeski, Paul Nassar, Noble Briggs, Jacob Hampton, Cathy Fairfield

**University of Miami, Miami, Florida**
Chris Mallow, Carolina Rivas

**University of Michigan, Ann Arbor, Michigan**
Weronika Valvano, Anne Kaniclades, Aleda Leis, Rebecca Fong, Mildred Wallace, Chiraag Balsara, Rachel Truson, Regina Lehmann, Weronika Damek Valvano, Abigail Carolan, Izza Imran, Jozyan Ujmaya

**University of Utah, Salt Lake City, Utah**
Bryce Bosworth, Amanda Orme

**University of Washington, Seattle, Washington**
Nicholas Johnson, Joshua Acidera, Maile McKeown, Dylan Clark, Leenay Coughlin, Frances Nagore

**Vanderbilt University Medical Center, Nashville, Tennessee**
Wesley Self, Yuwei Zhu, Cassandra Johnson, Adrienne Baughman, James D. Chappell, Natasha Halasa, Carlos G. Grijalva, Paul W. Blair, Jonathan D. Casey, Karen F. Miller, Jakea Johnson, Ian D. Jones, Kelsey N. Womack, Jillian Rhoads, Colleen Ratcliff, Sydney Cornelison,

Ine Sohn, Cara Lwin, Laura L. Short, Rendie E. McHenry, Jennifer L. Luther, Julio Angulo, Marcia Blair,  Shanice Cummings, Lauren Ezell, Emma Claire Gauthier, Samarian Hargrave, Anna Jackson, Jennifer Luther, Rendie McHenry, Bryan Peterson, Claudia Guevara Pulido, Neekar Rashid, Wanderson Rezende, Caroline Rice, Laura Short, Margaret Whitsett

**Wake Forest University, Winston-Salem, North Carolina**
Kevin Gibbs, Hannah Strait

**Washington University, St. Louis, Missouri**
Jennie Kwon, Bijal Parikh, David McDonald, Carleigh Samuels, Lucy Vogt, Caroline O'Neil, Alyssa Valencia, Francesca Yerbic, Olivia Arter, Kim Vu, Akshay Saluja, Elianora Ovchiyan, Sachina Mensah

**Yale University, New Haven, Connecticut**
Anirudh Goyal, Ivan Valesquez, Arda Yigitkanli, Kimberly Manchester, Lauren Delamielleure, Uchechi Okoronkwo

### Supplementary Methods

*Eligibility criteria and enrollment practices*

The Investigating Respiratory Viruses in the Acutely Ill (IVY) network is a multicenter inpatient surveillance network comprising 26 hospitals in 20 U.S. states used for assessing vaccine effectiveness and clinical epidemiology. Patients were enrolled according to the eligibility criteria listed below. Enrollment teams attempted to enroll all patients with laboratory-confirmed RSV based on clinical viral testing conducted at the enrolling hospital, if they met syndromic criteria for acute respiratory illness (see inclusion criteria below). In addition to clinical viral testing in the local hospital, nasal swabs were collected from all enrolled patients and systematically tested at a central laboratory (Vanderbilt University Medical Center) for RSV, SARS-CoV-2, and influenza by real-time reverse transcription-polymerase chain reaction (RT-PCR) using standardized methods.

*Enrollment inclusion criteria*

1. Age ≥18 years old.

2. Hospital admission or in an emergency department awaiting hospital admission.

3. Symptoms and/or signs compatible with an acute respiratory illness, including at least 1 of the following: fever; cough; shortness of breath; hypoxemia (for patients not on chronic supplemental oxygen, hypoxemia is defined as: $SpO_2 < 92\%$ or use of supplemental oxygen to maintain $SpO_2 \geq 92\%$; for patients on chronic supplemental oxygen, hypoxemia is defined as $SpO_2$ below the patient's baseline $SpO_2$ or an escalation of supplemental oxygen use to maintain the baseline $SpO_2$ value); new pulmonary findings on chest imaging consistent with pneumonia.

4. Clinically obtained test that is positive (for RSV cases) or negative (for test-negative control-patients) for acute RSV after onset of symptoms for the current illness. The test may be obtained before or after hospital arrival. Examples of acute viral tests include RT-PCR tests, other nucleic acid amplification tests (NAAT), and antigen tests. Serology testing may not be used for eligibility.

*Enrollment exclusion criteria*

1. Patient was admitted to the hospital more than 7 days ago (based on this exclusion criterion, patients must be enrolled within 7 days of hospital admission).

2. The first RSV test is known to have occurred more than 10 days after onset of acute viral infection symptoms/signs listed in inclusion criterion #3. Patients with unknown symptoms/signs onset date may be enrolled.

3. First test for acute RSV is more than 3 days after hospital admission.

4. Previously enrolled in this surveillance program within the prior 30 days.

*Analytic cohort*

Enrolled inpatients were included if they had RSV infection confirmed by clinical or central laboratory testing of a respiratory specimen collected within 10 days of symptom onset and 3 days of hospital admission. Patients with coinfection who also tested positive for SARS-CoV-2 or influenza, either locally based on clinical test results available at the time of enrollment or centrally in the IVY Network central laboratory, were excluded from this analysis.

*Classification of RSV vaccination status*

Vaccination status for RSV was determined from electronic medical records (EMR), state or jurisdictional registries, and by self- (or proxy-) report. Available vaccination data from each of these sources were collected, including date and location of vaccine administration, vaccine manufacturer, and lot number. Final vaccination status was determined by combining data from verified documented sources (EMR and registry data) as well as plausible self- (or proxy-) report based on date and location of RSV vaccination.

During the period of this analysis, the only two licensed and recommended RSV vaccine products were: 1) Recombinant RSVPreF3 adjuvanted (Arexvy, GlaxoSmithKline) and 2) Recombinant RSVPreF (Abrysvo, Pfizer, Inc.). RSV vaccination status was classified into three groups: 1) unvaccinated, defined as no receipt of RSV vaccine ever, 2) partially vaccinated, if they had received RSV vaccine <14 days from illness onset (these patients were excluded from the analysis); and 3) vaccinated, if they had received RSV vaccine ≥14 days before illness onset.

*Severe in-hospital outcomes*

Clinical severity of patients with RSV disease was characterized using the following severe in-hospital outcomes occurring from hospital presentation to hospital discharge, patient death, or hospital day 28:

i) RSV-associated intensive care unit (ICU) admission
ii) RSV-associated acute organ failure
iii) RSV-associated invasive mechanical ventilation (IMV) or death

<u>i) RSV-associated intensive care unit (ICU) admission</u>

Patients were classified as having RSV-associated ICU admission if they received care in an ICU for any duration of time during the hospitalization through day 28.

ii) RSV-associated acute organ failure

Patients were assessed for organ support therapies for the respiratory, cardiovascular, and renal systems during the index hospitalization through hospital day 28. Patients who were newly treated with any of the following organ support therapies met the definition for the acute organ failure outcome:

Organ support for respiratory failure: Patient who meets the definition for RSV-associated hospitalization plus receipt of high-flow nasal cannula (HFNC), non-invasive ventilation (NIV), or IMV during the index hospitalization before day 28. HFNC was defined as a supplemental oxygen flow rate of at least 30 liters per minute. NIV included both continuous positive airway pressure (CPAP) and bilevel positive airway pressure (BiPAP) delivered through a mask. A patient was classified as having NIV use in the hospital if NIV was received for therapy of the acute illness and not only for treatment of sleep apnea. IMV was defined as positive pressure administered through an endotracheal tube or tracheostomy tube. Patients who had chronic NIV use before the acute illness met the definition for respiratory virus-associated respiratory support if they had escalation of respiratory support to IMV in the hospital. Patients who had chronic IMV use prior to the acute illness were not eligible for the RSV-associated organ support for respiratory failure outcome.

Organ support for cardiovascular failure: Patient who meets the definition for RSV-associated hospitalization plus receipt of intravenous administration of a vasopressor medication by continuous infusion for any duration of time during the index hospitalization before day 28. Vasopressor medications included: norepinephrine, epinephrine, dopamine, phenylephrine, and vasopressin.

Organ support for renal failure: Patient who meets the definition for RSV-associated hospitalization plus receipt of new kidney replacement therapy during the index hospitalization before day 28. Any type of kidney replacement therapy would fulfill the definition for this outcome, including hemodialysis and continuous veno-venous hemofiltration. Patients with chronic kidney replacement therapy prior to the acute illness were not eligible for the RSV-associated organ support for renal failure outcome.

iii) RSV-associated IMV or death

Patients were classified as having RSV-associated IMV or death if they received IMV or died during the hospitalization through day 28. IMV was defined as positive pressure administered through an endotracheal tube or tracheostomy tube. Patients on home IMV prior to the acute illness could not meet the RSV-associated IMV or death outcome through receipt of in-hospital IMV.

*RSV detection by RT-PCR*

Total nucleic acid extract from 100 µl of upper respiratory specimen collected in viral transport medium was prepared using the MagNA Pure LC Total Nucleic Acid Isolation Kit (Roche Molecular Systems, Pleasanton, CA) and MagNA Pure 96 automated extraction platform (Roche) or QiaCube HT automated extraction system (Qiagen, Germantown, MD) and QIAamp 96 Virus QiaCube HT kit (Qiagen). Extracts (100 µl eluate volume) were tested by RT-PCR on the StepOnePlus, QuantStudio 3, QuantStudio 5, or QuantStudio 6 Real-Time PCR System (Applied Biosystems, Waltham, MA) for a pan-RSV matrix gene target using Superscript III Platinum One-Step Quantitative RT-PCR System containing ROX passive reference dye (Invitrogen, Waltham, MA) and a screening set of primers and probe (forward: GGCAAATATGGAAACATACGTGAA; reverse: TCTTTTTCTAGGACATTGTAYTGAACAG; probe: FAM-CTGTGTATGTGGAGCCTTCGTGAAGCT-BHQ-1) (Biosearch Technologies, Petaluma, CA). Subgroup differentiation of RSV screen-positive specimens was performed by RT-PCR using Superscript III, a common set of primers, and unique probes targeting A- and B-specific sequences in the viral polymerase (L) gene (forward: AATACAGCCAAATCTAACCAACTTTACA; reverse: GCCAAGGAAGCATGCAATAAA; RSV-A probe: FAM-TGCTATTGTGCACTAAAG-MGBNFQ; RSV-B probe: VIC-CACTATTCCTTACTAAAGATGTC-MGBNFQ) (Thermo Fisher, Waltham, MA). Each specimen also was tested for human RNase P (RNP) gene sequence as a marker of specimen adequacy and sensor for PCR inhibitors using TaqPath 1-Step RT-qPCR Master Mix, CG (Applied Biosystems). PCR reactions consisted of 45 amplification cycles, and Ct values of any magnitude were accepted when represented by a characteristic specific amplification curve. Valid A or B subgroup identification was contingent on co-detection of the universal RSV target. Specimens generating unrepeatable weakly positive signal in the screening assay and negative results in both subgrouping assays were considered inconclusive for viral RNA. Absence of RSV detection in specimens registering RNP Ct values ≥40 was considered inconclusive for viral RNA.

*Selected underlying conditions*

We included underlying conditions potentially related to severe RSV disease as defined by Britton et al. [1]. Underlying medical conditions were obtained through medical record review. We used individual underlying conditions when possible as indicator (input) variables without collapsing conditions into broad organ system categories. Collapsing conditions into an organ system category could discard information on both severity (e.g., a patient presenting with mild asthma is treated equivalently to a patient with severe COPD) and the number of symptoms occurring within organ system groups. Additionally, organ system groupings are not distinct boundaries, and symptoms can overlap multiple organ systems.

To restrict to the most informative conditions, we removed variables with a high percentage (≥10%) of missing data (n=0), moderate correlation with other indicator variables (as defined by tetrachoric correlation ≥0.7) (n=1; neuromuscular conditions and feeding tube use, of which neuromuscular conditions was retained), and rare (≤1%) prevalence (n=2; sickle cell disease or thalassemia among adults aged ≥60 and dementia among adults aged 18–59 were excluded). We used tetrachoric correlation as most clinical variables we observed were binary with some variables potentially representing an underlying continuous gradient of disease severity, in line with assumptions underlying this method [2]. Results were qualitatively similar to those obtained using Pearson correlation, but the magnitudes of Pearson correlation tended to be lower.

Additionally, we created four composite categorical variables to measure granularity in condition severity:

- Chronic obstructive pulmonary disease (COPD) stratified by home oxygen use (levels: no COPD, COPD without home oxygen use, COPD with home oxygen use)
    - Home oxygen use was defined as any home oxygen use including home non-invasive and invasive ventilation
- Other pulmonary disease (defined as any of cystic fibrosis, pulmonary fibrosis, or pulmonary hypertension) stratified by stratified by home oxygen use (levels: no other pulmonary disease, pulmonary disease without home oxygen use, pulmonary disease with home oxygen use)
- Liver disease (levels: no liver disease, liver disease without cirrhosis, liver disease with cirrhosis)
- Chronic kidney disease (levels: no chronic kidney disease, chronic kidney disease without chronic renal replacement therapy, end stage renal disease on chronic renal replacement therapy)

This resulted in the following 17 variables in total, with 16 for each age group:

- BMI categories as defined using the following BMI (kg/$m^2$) ranges: underweight (less than 18.5); healthy weight (18.5 to less than 25); overweight (25 to less than 30); class 1 or 2 obesity (30 to less than 40); class 3 i.e. severe obesity (40 or greater). For counts of underlying conditions, only severe obesity was included among all BMI categories to align with vaccine eligibility guidelines [1], but for regression models, all BMI categories were included.
- Heart failure
- Atherosclerotic cardiovascular disease (atherosclerotic CVD; defined as any prior myocardial infection, peripheral vascular disease, prior stroke, or prior transient ischemic attack)
- Chronic obstructive pulmonary disease (COPD) stratified by home oxygen use
- Asthma
- Other pulmonary disease (defined as any of cystic fibrosis, pulmonary fibrosis, or pulmonary hypertension) stratified by home oxygen use
- Dementia (only included for adults aged ≥60)
- Neuromuscular conditions (defined as any of muscular dystrophy, cerebral palsy, hemiplegia, paraplegia, or anterolateral sclerosis)
- Liver disease (levels: no liver disease, liver disease without cirrhosis, liver disease with cirrhosis)
- Diabetes

- Chronic kidney disease (CKD; levels: no chronic kidney disease, chronic kidney disease without chronic renal replacement therapy, end stage renal disease on chronic renal replacement therapy)
- Hemoglobinopathies (sickle cell disease or thalassemia; only included for adults aged 18–59)
- Autoimmune or autoinflammatory condition (defined as any of systemic lupus erythematosus, rheumatoid arthritis, psoriasis, polymyositis, mixed connective tissue disease, polymyalgia rheumatica, inflammatory bowel disease, or other autoimmune diseases); these conditions were grouped as the prevalence of each individual condition was low
- HIV infection
- Active solid tumor malignancy
- Active hematologic malignancy
- Solid organ or hematopoietic cell transplant recipient

*Bayesian profile regression model*

This analysis used Bayesian profile regression, an outcome-informed statistical model developed to address limitations of traditional regression methods, which may be difficult to fit or interpret when underlying conditions are highly correlated [3,4]. Statistical clustering approaches have been applied successfully in other disease contexts to parse correlated symptoms or underlying conditions data into more interpretable clinical groupings [5–10]. For instance, latent class analysis of underlying conditions among 12,340 decedents with COVID-19 patients identified three clusters of conditions, comprising cardiovascular disease and diabetes, cardiovascular disease without diabetes, and a minimal prevalence class [11]. Bayesian profile regression and similar methods identify probabilistic profiles or clusters with distinct underlying condition prevalences, and can generate hypotheses for developing interpretable and formally defined clinical subgroups or risk scores.

Extension of latent class analysis and related approaches to consider information from outcome variables is an active area of investigation [12], and is a strength of the outcome-informed Bayesian profile regression approach we have used here [3,13,14]. Bayesian profile regression comprises an *assignment submodel* (assigns individuals to a condition profile; modeled using a Dirichlet Process mixture model) and a *disease submodel* (evaluates association of profiles with the outcome variable; modeled using a regression framework) which are jointly fit [3,13–15].

For an individual $i$ with underlying conditions data $X_i$ and dichotomous severe outcome $Y_i$, the joint likelihood for these two submodels assuming profile membership is known is given by:

$$P(X_i, Y_i \mid Z_i = c, \theta_c, \Phi_c) = P(X_i \mid Z_i = c, \Phi_c)\, P(Y_i \mid Z_i = c, \theta_c)$$

Where $Z_i$ is a latent profile allocation variable indicating that individual $i$ belongs to profile $c$, $\theta_c$ measures the effect of profile $c$ on the severe outcome variable, and $\Phi_c$ describes profile-specific probabilities of the occurrence of the $J$ conditions. Assuming conditions are categorical, we can denote $K_j$ as the number of levels in condition $j$ (typically two as most conditions are dichotomous); $\Phi_c$ is given by:

$$\Phi_c = [\Phi_{c,1}, \dots, \Phi_{c,J}]$$

$$\Phi_{c,j} = [\phi_{c,j,1}, \dots, \phi_{c,j,K_j}]$$

The $\Phi_c$ vector can be interpreted as the average condition probability within a profile, allowing profiles to be distinguished by conditions with particularly high or low prevalence (see below). With $X_{ij}$ representing the $j$th condition for individual $i$, the *assignment submodel* can then be written as:

$$P(X_i \mid Z_i = c, \Phi_c) = \Pi_{j=1}^{J} \phi_{c,j,X_{ij}}$$

This represents the likelihood of observing an individual's underlying conditions given membership in a profile. Note that conditional independence (underlying conditions are independent given profile membership) is assumed, similar to as in latent class analysis and other discrete mixture models [16,17]. In latent class analysis, unmodeled conditional dependence can lead to biased estimates and increased misclassification [18].

Allowing for an infinite number of profiles $c \in [1, 2, \dots]$, the mixture weights for profile membership are defined as $\boldsymbol{\psi}$, where $P(Z_i = c \mid \boldsymbol{\psi}) = \psi_c$. $Z_i \mid \boldsymbol{\psi}$ is constructed using the stick-breaking formulation of the Dirichlet process (see [13]).

The *disease submodel* defines the assigned profile to be a predictor of the severe outcome, directly allowing the outcome variable to influence profile assignments. We use a logit link function for the binary outcome variable:

$$P(Y_i = 1 \mid Z_i = c, \theta_c) = logit^{-1}(\theta_c) = \frac{1}{1 + e^{-\theta_c}}$$

In this disease submodel, we have not included any additional fixed effect covariates. We have partially accounted for the effect of age by conducting an age-stratified analysis (see Discussion for limitations).

Now considering the full cohort, we denote $\boldsymbol{Z} = [Z_1, \dots, Z_n]$ and similarly for $\boldsymbol{X}, \boldsymbol{Y}$. With profile membership index $c \in [1, 2, \dots]$, we then denote $\boldsymbol{\theta} = [\theta_1, \theta_2, \dots, \theta_c]$ and similarly for $\boldsymbol{\psi}$ and $\boldsymbol{\Phi}$. The full posterior distribution is given by:

$$P(\boldsymbol{Z}, \boldsymbol{\psi}, \boldsymbol{\theta}, \boldsymbol{\Phi}, \alpha \mid \boldsymbol{X}, \boldsymbol{Y}) \propto P(\boldsymbol{X}, \boldsymbol{Y} \mid \boldsymbol{Z}, \boldsymbol{\theta}, \boldsymbol{\Phi})\, P(\boldsymbol{Z} \mid \boldsymbol{\psi})\, P(\boldsymbol{\psi} \mid \alpha)\, P(\boldsymbol{\theta})\, P(\boldsymbol{\Phi})$$

The prior distributions are described below along with default hyperparameters (see [3,13] for detail):

- The mixture weights given by $\boldsymbol{\psi}$ follow a Dirichlet Process prior distribution where $P(\alpha)$ is modeled by a Gamma distribution with shape = 2 and scale = 1.
- The $\theta_c$ variables are assumed to each follow a *t*-distribution with 7 degrees of freedom.
- The $\Phi_{c,j}$ variables are assumed to each follow a Dirichlet prior distribution with hyperparameter $a_j$ set to 1 for all conditions.

The posterior distribution is sampled using the Markov chain Monte Carlo (MCMC) approach described by Liverani et al. and implemented in the R PReMiuM package [13].

*Model fitting, post-processing, and sensitivity analysis*

We ran PReMiuM (version 3.2.13) with 5,000 burn-in (i.e., discarded) iterations, 10,000 analytic iterations after burn-in, 15 initial profiles, and default hyperparameters. Trace plots for $\alpha$ and the number of profiles indicated no evidence against convergence. We then conducted Molitor et al.'s postprocessing approach for identifying a "typical" partition from the stochastic results of sampling from the posterior distribution [3]. A pairwise similarity matrix was first calculated representing the degree of co-clustering of patients across the 10,000 MCMC iterations. Partitioning around medoids was then used to construct an optimal partitioning consistent with the similarity matrix after maximizing the average silhouette width, a measure of clustering quality [3]. One advantage of this postprocessing approach is that the width of the credible intervals will also reflect uncertainty from the profile assignment process [3].

To assess dependency of MCMC and this postprocessing method on initial random profile allocations and different severe in-hospital outcomes, we re-ran the models for all three outcomes 25 times using different initial allocations. Among adults aged 18–59, two profiles were supported in all 25 iterations for all three severe in-hospital outcomes. Among adults aged ≥60, four profiles were supported in the majority of iterations for ICU admission (24/25 iterations), acute organ failure (24/25), and IMV or death (16/25). We used the number of profiles (4) supported by the majority of iterations. Profile membership across all three outcomes in both age groups were highly similar as assessed by the Adjusted Rand Index (>0.8), and we selected one iteration of an ICU admission model for defining profile membership.

We calculated median posterior and 95% equal-tailed credible intervals (i.e., spanning from the $2.5^{th}$ to $97.5^{th}$ percentile of the posterior distribution) for $\Phi_c$ and $P(Y_i = 1 \mid \theta_c)$ to summarize the posterior distribution using the optimal partition identified above. We interpreted profile membership based on profile-defining conditions where credible intervals for $\phi_{c,j}$ excluded overall averages. To evaluate differences in risk between profiles directly, we calculated risk ratios $RR_{Profile} = P(Y_i = 1 \mid \theta_c) \ / \ P(Y_i = 1 \mid \theta_1)$ relative to patients in the minimal prevalence (defined in Results) profile within each age group. We interpreted $RR_{Profile}$ where credible intervals excluded 1 as evidence for a change in risk of the severe in-hospital outcome relative to the reference (minimal prevalence) profile. We also calculated the posterior probability $P(RR_{Profile} > 1)$ to quantify the strength of evidence for increased risk.

*Poisson regression*

We used Poisson regression models to estimate risk ratios ($RR_{Poisson}$) for severe in-hospital outcomes among patients with versus without underlying conditions. Modified Poisson regression (i.e., with robust error variance) has been previously proposed as a method to estimate RRs [19] and has been suggested to perform better under model misspecification compared to log-binomial regression [20]. As several underlying conditions were rare, we additionally included a Firth-type penalty and the Morel correction for calculating robust error variances using the *firthb* R package as described and implemented by Uno et al. [21].

**Supplementary Table 1. Characteristics and Number of Underlying Conditions[a] Associated with Respiratory Syncytial Virus Disease Among Bayesian Profiles[b] of 397 Patients Aged 18–59 Hospitalized with Respiratory Syncytial Virus — IVY Network, 26 Hospitals, January 2022–July 2024**

| | Overall (N=397) | Profile 1 (N=308) | Profile 2 (N=89) |
|---:|---:|---:|---:|
| **Age** | | | |
| Median (IQR) | 49 (35, 55) | 48 (34, 54) | 51 (46, 56) |
| **Sex** | | | |
| Female | 213 (53.7%) | 171 (55.5%) | 42 (47.2%) |
| Male | 184 (46.3%) | 137 (44.5%) | 47 (52.8%) |
| **Race/ethnicity** | | | |
| Hispanic | 77 (19.4%) | 64 (20.8%) | 13 (14.6%) |
| Non-Hispanic Black | 122 (30.7%) | 89 (28.9%) | 33 (37.1%) |
| Non-Hispanic White | 161 (40.6%) | 125 (40.6%) | 36 (40.4%) |
| Non-Hispanic, other race[c] | 21 (5.3%) | 14 (4.5%) | 7 (7.9%) |
| Other[d] | 16 (4.0%) | 16 (5.2%) | 0 (0%) |
| **Ribavirin administered[e]** | 20 (5.9%) | 17 (6.5%) | 3 (3.9%) |
| **BMI category[f]** | | | |
| Underweight | 20 (5.0%) | 13 (4.2%) | 7 (7.9%) |
| Healthy weight | 98 (24.7%) | 80 (26.0%) | 18 (20.2%) |
| Overweight | 81 (20.4%) | 65 (21.1%) | 16 (18.0%) |
| Obesity | 131 (33.0%) | 99 (32.1%) | 32 (36.0%) |
| Severe obesity | 67 (16.9%) | 51 (16.6%) | 16 (18.0%) |
| **Average number of underlying conditions** | | | |
| Median (IQR) | 2.0 (1.0, 3.0) | 1.0 (1.0, 2.0) | 3.0 (3.0, 4.0) |
| **Number of underlying conditions** | | | |
| 0 | 51 (12.8%) | 51 (16.6%) | 0 (0%) |
| 1 | 122 (30.7%) | 121 (39.3%) | 1 (1.1%) |
| 2 | 95 (23.9%) | 77 (25.0%) | 18 (20.2%) |
| 3 | 77 (19.4%) | 44 (14.3%) | 33 (37.1%) |
| ≥4 | 52 (13.1%) | 15 (4.9%) | 37 (41.6%) |
| **ICU admission** | 96 (24.2%) | 56 (18.2%) | 40 (44.9%) |
| **Acute organ failure[g]** | 118 (29.7%) | 78 (25.3%) | 40 (44.9%) |
| **IMV or death** | 32 (8.1%) | 20 (6.5%) | 12 (13.5%) |

| | Overall (N=397) | Profile 1 (N=308) | Profile 2 (N=89) |
|---|---|---|---|
| **Days from symptom onset to admission[h]** Median (IQR) | 3.0 (1.0, 4.0) | 2.0 (1.0, 4.0) | 3.0 (2.0, 6.0) |

Percentages are column percentages. Among adults aged 18–59, profile 1 represents the "minimal prevalence" profile and 2 represents the "cardiorenal and diabetes" profile.

[a] For counts of underlying conditions, only severe obesity was included among all BMI categories to align with vaccine eligibility guidelines [1]. Other underlying conditions associated with RSV severe disease and included in counts were diabetes, chronic kidney disease (including end-stage renal disease), heart failure, atherosclerotic cardiovascular disease, asthma, chronic obstructive pulmonary disease (with or without home oxygen use), other pulmonary conditions (cystic fibrosis, pulmonary fibrosis, or pulmonary hypertension; with or without home oxygen use), solid organ or hematopoietic cell transplant receipt, active hematologic malignancy, active solid tumor malignancy, HIV infection, autoimmune or autoinflammatory conditions (systemic lupus erythematosus, rheumatoid arthritis, psoriasis, polymyositis, mixed connective tissue disease, polymyalgia rheumatica, inflammatory bowel disease, or other autoimmune diseases), dementia (only included for adults aged ≥60), neuromuscular conditions (muscular dystrophy, cerebral palsy, hemiplegia, paraplegia, or anterolateral sclerosis), liver disease (including cirrhosis), and sickle cell disease or thalassemia (only included for adults aged 18–59).

[b] Bayesian profile regression is an outcome variable-informed statistical clustering approach that comprises an assignment submodel (assigns individuals to a condition profile) and a disease submodel (evaluates association of profiles with the outcome variable); the two submodels are jointly fitted in a Bayesian paradigm, enabling the outcome variable to influence profile membership.

[c] "Non-Hispanic, other race" includes American Indian or Alaska Native, Asian, and Native Hawaiian or other Pacific Islander categories, which were combined because of small counts.

[d] "Other" includes patients who self-reported their race and ethnicity as "other" and those for whom race and ethnicity were unknown.

[e] A total of 58 patients (14.6%) were missing data on ribavirin use. The percentage of patients receiving ribavirin was calculated based on nonmissing data.

[f] BMI categories were defined using the following BMI (kg/$m^2$) ranges: underweight (less than 18.5); healthy weight (18.5 to less than 25); overweight (25 to less than 30); class 1 or 2 obesity (30 to less than 40); class 3 i.e. severe obesity (40 or greater).

[g] Acute organ failure defined as a composite of respiratory failure (new receipt of high-flow nasal canula, non-invasive mechanical ventilation, or IMV), cardiovascular failure (use of vasopressors), or kidney failure (new receipt of kidney replacement therapy).

[h] A total of 114 patients (28.7%) were missing data on symptom onset date. The median time between symptom onset and hospital admission was calculated based on nonmissing data.

Abbreviations: RSV = respiratory syncytial virus; ICU = intensive care unit; BMI = body mass index; IMV = intensive mechanical ventilation.

**Supplementary Table 2. Prevalence of Underlying Conditions Associated with Respiratory Syncytial Virus Disease Among Bayesian Profiles[a] of 397 Patients Aged 18–59 Hospitalized with Respiratory Syncytial Virus — IVY Network, 26 Hospitals, January 2022–July 2024**

| | Overall (N=397) | 1 (N=308) | 2 (N=89) | P-value |
|---|---|---|---|---|
| **BMI category** | | | | 0.484 |
| Underweight | 20 (5.0%) | 13 (4.2%) | 7 (7.9%) | |
| Healthy weight | 98 (24.7%) | 80 (26.0%) | 18 (20.2%) | |
| Overweight | 81 (20.4%) | 65 (21.1%) | 16 (18.0%) | |
| Obesity | 131 (33.0%) | 99 (32.1%) | 32 (36.0%) | |
| Severe obesity | 67 (16.9%) | 51 (16.6%) | 16 (18.0%) | |
| **Diabetes** | | | | <0.0001 |
| No | 288 (72.5%) | 242 (78.6%) | 46 (51.7%) | |
| Yes | 109 (27.5%) | 66 (21.4%) | 43 (48.3%) | |
| **Chronic kidney disease** | | | | <0.0001 |
| No | 310 (78.1%) | 286 (92.9%) | 24 (27.0%) | |
| Yes; ESRD | 45 (11.3%) | 5 (1.6%) | 40 (44.9%) | |
| Yes; without ESRD | 42 (10.6%) | 17 (5.5%) | 25 (28.1%) | |
| **Heart failure** | | | | <0.0001 |
| No | 314 (79.1%) | 282 (91.6%) | 32 (36.0%) | |
| Yes | 83 (20.9%) | 26 (8.4%) | 57 (64.0%) | |
| **Atherosclerotic CVD** | | | | <0.0001 |
| No | 351 (88.4%) | 290 (94.2%) | 61 (68.5%) | |
| Yes | 46 (11.6%) | 18 (5.8%) | 28 (31.5%) | |
| **Asthma** | | | | <0.0001 |
| No | 301 (75.8%) | 218 (70.8%) | 83 (93.3%) | |
| Yes | 96 (24.2%) | 90 (29.2%) | 6 (6.7%) | |
| **COPD** | | | | <0.0001 |
| No | 345 (86.9%) | 281 (91.2%) | 64 (71.9%) | |
| Yes; home O2 use | 12 (3.0%) | 3 (1.0%) | 9 (10.1%) | |
| Yes; no home O2 use | 40 (10.1%) | 24 (7.8%) | 16 (18.0%) | |
| **Other pulmonary** | | | | <0.0001 |
| No | 364 (91.7%) | 292 (94.8%) | 72 (80.9%) | |
| Yes; home O2 use | 9 (2.3%) | 2 (0.6%) | 7 (7.9%) | |
| Yes; no home O2 use | 24 (6.0%) | 14 (4.5%) | 10 (11.2%) | |

| | Overall (N=397) | 1 (N=308) | 2 (N=89) | P-value |
|---|---|---|---|---|
| **Transplant receipt** | | | | <0.0001 |
| No | 338 (85.1%) | 282 (91.6%) | 56 (62.9%) | |
| Yes | 59 (14.9%) | 26 (8.4%) | 33 (37.1%) | |
| **Hematalogic malignancy** | | | | 0.408 |
| No | 364 (91.7%) | 280 (90.9%) | 84 (94.4%) | |
| Yes | 33 (8.3%) | 28 (9.1%) | 5 (5.6%) | |
| **Solid tumor** | | | | 0.0846 |
| No | 376 (94.7%) | 288 (93.5%) | 88 (98.9%) | |
| Yes | 21 (5.3%) | 20 (6.5%) | 1 (1.1%) | |
| **HIV** | | | | 0.146 |
| No | 373 (94.0%) | 286 (92.9%) | 87 (97.8%) | |
| Yes | 24 (6.0%) | 22 (7.1%) | 2 (2.2%) | |
| **Autoimmune or IBD** | | | | 1 |
| No | 363 (91.4%) | 282 (91.6%) | 81 (91.0%) | |
| Yes | 34 (8.6%) | 26 (8.4%) | 8 (9.0%) | |
| **Neuromuscular disease** | | | | 0.0852 |
| No | 383 (96.5%) | 294 (95.5%) | 89 (100%) | |
| Yes | 14 (3.5%) | 14 (4.5%) | 0 (0%) | |
| **Liver disease** | | | | 0.264 |
| No | 382 (96.2%) | 298 (96.8%) | 84 (94.4%) | |
| Yes; cirrhosis | 9 (2.3%) | 5 (1.6%) | 4 (4.5%) | |
| Yes; without cirrhosis | 6 (1.5%) | 5 (1.6%) | 1 (1.1%) | |
| **Sickle cell or thalassemia** | | | | 0.328 |
| No | 390 (98.2%) | 301 (97.7%) | 89 (100%) | |
| Yes | 7 (1.8%) | 7 (2.3%) | 0 (0%) | |

Percentages are column percentages. *P*-values are calculated using the Chi-square test. Among adults aged 18–59, profile 1 represents the “minimal prevalence” profile and 2 represents the “cardiorenal and diabetes” profile.

[a] Bayesian profile regression is an outcome variable-informed statistical clustering approach that comprises an assignment submodel (assigns individuals to a condition profile) and a disease submodel (evaluates association of profiles with the outcome variable); the two submodels are jointly fitted in a Bayesian paradigm, enabling the outcome variable to influence profile membership.

Abbreviations: RSV = respiratory syncytial virus; BMI = body mass index.

**Supplementary Table 3. Prevalence of Heart Failure, Chronic Kidney Disease, and Diabetes Among Bayesian Cardiorenal and Diabetes Profile[a] of Patients Hospitalized with Respiratory Syncytial Virus by Age Group — IVY Network, 26 Hospitals, January 2022–July 2024**

| **Patients aged 18–59 (N = 89)** | | | | | |
|---|---|---|---|---|---|
| **Heart failure** | Chronic kidney disease (including ESRD) | Diabetes | N | Prevalence (%) | Number of conditions |
| **Yes** | Yes | Yes | 16 | 18 | 3 |
| **No** | Yes | Yes | 15 | 16.9 | 2 |
| **Yes** | No | Yes | 10 | 11.2 | 2 |
| **Yes** | Yes | No | 22 | 24.7 | 2 |
| **No** | No | Yes | 2 | 2.2 | 1 |
| **No** | Yes | No | 12 | 13.5 | 1 |
| **Yes** | No | No | 9 | 10.1 | 1 |
| **No** | No | No | 3 | 3.4 | 0 |
| **Patients aged ≥60 (N = 183)** | | | | | |
| **Heart failure** | Chronic kidney disease (including ESRD) | Diabetes | N | Prevalence (%) | Number of conditions |
| **Yes** | Yes | Yes | 57 | 31.1 | 3 |
| **No** | Yes | Yes | 46 | 25.1 | 2 |
| **Yes** | No | Yes | 25 | 13.7 | 2 |
| **Yes** | Yes | No | 32 | 17.5 | 2 |
| **No** | No | Yes | 1 | 0.5 | 1 |
| **No** | Yes | No | 15 | 8.2 | 1 |
| **Yes** | No | No | 7 | 3.8 | 1 |

[a] Bayesian profile regression is an outcome variable-informed statistical clustering approach that comprises an assignment submodel (assigns individuals to a condition profile) and a disease submodel (evaluates association of profiles with the outcome variable); the two submodels are jointly fitted in a Bayesian paradigm, enabling the outcome variable to influence profile membership.

**Supplementary Table 4. Risk Ratios and Posterior Probabilities of Severe Outcomes[a] Among Bayesian Profiles[b] of Patients Hospitalized with Respiratory Syncytial Virus — IVY Network, 26 Hospitals, January 2022–July 2024**

| Age group | Outcome | Profile | $RR_{Profile}$ [95% credible interval] | $P(RR_{Profile} > 1)$ |
|---|---|---|---|---|
| 18–59 | ICU admission | 2 | 1.81 [1.26–2.56] * | 0.9996 |
| 18–59 | Acute organ failure | 2 | 1.37 [1.00–1.82] * | 0.9746 |
| 18–59 | Death or IMV | 2 | 1.51 [0.70–2.88] | 0.8685 |
| ≥60 | ICU admission | 2 | 1.26 [0.96–1.62] | 0.9456 |
| ≥60 | ICU admission | 3 | 0.55 [0.26–0.98] * | 0.0221 |
| ≥60 | ICU admission | 4 | 2.03 [1.11–3.12] * | 0.9870 |
| ≥60 | Acute organ failure | 2 | 1.25 [0.99–1.56] | 0.9710 |
| ≥60 | Acute organ failure | 3 | 0.77 [0.48–1.17] | 0.1172 |
| ≥60 | Acute organ failure | 4 | 1.90 [1.15–2.65] * | 0.9929 |
| ≥60 | Death or IMV | 2 | 2.00 [1.31–3.04] * | 0.9977 |
| ≥60 | Death or IMV | 3 | 1.23 [0.56–2.52] | 0.7042 |
| ≥60 | Death or IMV | 4 | 1.20 [0.13–3.88] | 0.5859 |

* Indicates 95% credible intervals that exclude the null.

Risk ratios ($RR_{Profile}$) were calculated using profile 1 (minimal prevalence profile) as the reference group within each age group, respectively. Point estimates are median posterior distribution risk ratio estimates. Among adults in both age groups, profile 1 represents the minimal prevalence profile and 2 represents the cardiorenal and diabetes profile. Among adults aged ≥60, profile 3 represents the hematologic malignancy profile, and 4 represents the severe chronic pulmonary disease with home oxygen dependence profile.

[a] Severe outcomes included ICU admission, acute organ failure, and invasive mechanical ventilation (IMV) or death. Acute organ failure was defined as a composite of respiratory failure (new receipt of high-flow nasal canula, non-invasive mechanical ventilation, or IMV), cardiovascular failure (use of vasopressors), or kidney failure (new receipt of kidney replacement therapy).

[b] Bayesian profile regression is an outcome variable-informed statistical clustering approach that comprises an assignment submodel (assigns individuals to a condition profile) and a disease

submodel (evaluates association of profiles with the outcome variable); the two submodels are jointly fitted in a Bayesian paradigm, enabling the outcome variable to influence profile membership.

Abbreviations: ICU = intensive care unit; IMV = intensive mechanical ventilation.

**Supplementary Table 5. Characteristics and Number of Underlying Conditions[a] Associated with Respiratory Syncytial Virus Disease Among Bayesian Profiles[b] of 714 Patients Aged ≥60 Hospitalized with Respiratory Syncytial Virus — IVY Network, 26 Hospitals, January 2022–July 2024**

| | Overall (N=714) | Profile 1 (N=460) | Profile 2 (N=183) | Profile 3 (N=52) | Profile 4 (N=19) |
|---|---|---|---|---|---|
| **Age** | | | | | |
| Median (IQR) | 72 (66, 80) | 73 (66, 81) | 73 (67, 80) | 70 (66, 75) | 71 (69, 82) |
| **Sex** | | | | | |
| Female | 387 (54.2%) | 255 (55.4%) | 99 (54.1%) | 23 (44.2%) | 10 (52.6%) |
| Male | 327 (45.8%) | 205 (44.6%) | 84 (45.9%) | 29 (55.8%) | 9 (47.4%) |
| **Race/ethnicity** | | | | | |
| Hispanic | 104 (14.6%) | 59 (12.8%) | 31 (16.9%) | 10 (19.2%) | 4 (21.1%) |
| Non-Hispanic Black | 139 (19.5%) | 83 (18.0%) | 44 (24.0%) | 8 (15.4%) | 4 (21.1%) |
| Non-Hispanic White | 415 (58.1%) | 287 (62.4%) | 87 (47.5%) | 31 (59.6%) | 10 (52.6%) |
| Non-Hispanic, other race[c] | 31 (4.3%) | 16 (3.5%) | 13 (7.1%) | 1 (1.9%) | 1 (5.3%) |
| Other[d] | 25 (3.5%) | 15 (3.3%) | 8 (4.4%) | 2 (3.8%) | 0 (0%) |
| **RSV vaccination[e]** | 10 (1.4%) | 4 (0.9%) | 4 (2.2%) | 0 (0%) | 2 (10.5%) |
| **Ribavirin administered[f]** | 26 (4.1%) | 3 (0.7%) | 7 (4.3%) | 16 (37.2%) | 0 (0%) |
| **BMI category[g]** | | | | | |
| Underweight | 53 (7.4%) | 42 (9.1%) | 9 (4.9%) | 1 (1.9%) | 1 (5.3%) |
| Healthy weight | 199 (27.9%) | 137 (29.8%) | 38 (20.8%) | 17 (32.7%) | 7 (36.8%) |
| Overweight | 210 (29.4%) | 144 (31.3%) | 43 (23.5%) | 21 (40.4%) | 2 (10.5%) |
| Obesity | 182 (25.5%) | 94 (20.4%) | 71 (38.8%) | 10 (19.2%) | 7 (36.8%) |
| Severe obesity | 70 (9.8%) | 43 (9.3%) | 22 (12.0%) | 3 (5.8%) | 2 (10.5%) |
| **Average number of underlying conditions** | | | | | |
| Median (IQR) | 2.0 (1.0, 3.0) | 1.0 (1.0, 2.0) | 4.0 (3.0, 4.0) | 3.0 (2.0, 4.0) | 4.0 (3.0, 5.0) |
| **Number of underlying conditions** | | | | | |
| 0 | 65 (9.1%) | 65 (14.1%) | 0 (0%) | 0 (0%) | 0 (0%) |
| 1 | 181 (25.4%) | 173 (37.6%) | 3 (1.6%) | 5 (9.6%) | 0 (0%) |
| 2 | 179 (25.1%) | 132 (28.7%) | 26 (14.2%) | 19 (36.5%) | 2 (10.5%) |
| 3 | 147 (20.6%) | 65 (14.1%) | 62 (33.9%) | 14 (26.9%) | 6 (31.6%) |
| ≥4 | 142 (19.9%) | 25 (5.4%) | 92 (50.3%) | 14 (26.9%) | 11 (57.9%) |

| | Overall (N=714) | Profile 1 (N=460) | Profile 2 (N=183) | Profile 3 (N=52) | Profile 4 (N=19) |
|---|---|---|---|---|---|
| **ICU admission** | 164 (23.0%) | 97 (21.1%) | 54 (29.5%) | 4 (7.7%) | 9 (47.4%) |
| **Acute organ failure[h]** | 225 (31.5%) | 138 (30.0%) | 67 (36.6%) | 10 (19.2%) | 10 (52.6%) |
| **IMV or death** | 70 (9.8%) | 36 (7.8%) | 28 (15.3%) | 4 (7.7%) | 2 (10.5%) |
| **Days from symptom onset to admission[i]** Median (IQR) | 2.0 (1.0, 4.0) | 3.0 (1.0, 4.0) | 2.0 (1.0, 4.0) | 3.0 (1.0, 6.0) | 2.0 (2.0, 4.0) |

Percentages are column percentages. Among adults aged ≥60, profile 1 represents the minimal prevalence profile, 2 represents the cardiorenal and diabetes profile, 3 represents the hematologic malignancy profile, and 4 represents the severe chronic pulmonary disease with home oxygen dependence profile.

[a] For counts of underlying conditions, only severe obesity was included among all BMI categories to align with vaccine eligibility guidelines [1]. Other underlying conditions associated with RSV severe disease and included in counts were diabetes, chronic kidney disease (including end-stage renal disease), heart failure, atherosclerotic cardiovascular disease, asthma, chronic obstructive pulmonary disease (with or without home oxygen use), other pulmonary conditions (cystic fibrosis, pulmonary fibrosis, or pulmonary hypertension; with or without home oxygen use), solid organ or hematopoietic cell transplant receipt, active hematologic malignancy, active solid tumor malignancy, HIV infection, autoimmune or autoinflammatory conditions (systemic lupus erythematosus, rheumatoid arthritis, psoriasis, polymyositis, mixed connective tissue disease, polymyalgia rheumatica, inflammatory bowel disease, or other autoimmune diseases), dementia (only included for adults aged ≥60), neuromuscular conditions (muscular dystrophy, cerebral palsy, hemiplegia, paraplegia, or anterolateral sclerosis), liver disease (including cirrhosis), and sickle cell disease or thalassemia (only included for adults aged 18–59).

[b] Bayesian profile regression is an outcome variable-informed statistical clustering approach that comprises an assignment submodel (assigns individuals to a condition profile) and a disease submodel (evaluates association of profiles with the outcome variable); the two submodels are jointly fitted in a Bayesian paradigm, enabling the outcome variable to influence profile membership.

[c] “Non-Hispanic, other race” includes American Indian or Alaska Native, Asian, and Native Hawaiian or other Pacific Islander categories, which were combined because of small counts.

[d] “Other” includes patients who self-reported their race and ethnicity as “other” and those for whom race and ethnicity were unknown.

[e] RSV vaccination status was determined from electronic medical records, immunization registries, and self-report and was defined as RSV vaccine receipt 14 days or more before illness onset.

[f] A total of 85 patients (11.9%) were missing data on ribavirin use. The percentage of patients receiving ribavirin was calculated based on nonmissing data.

[g] BMI categories were defined using the following BMI ($kg/m^2$) ranges: underweight (less than 18.5); healthy weight (18.5 to less than 25); overweight (25 to less than 30); class 1 or 2 obesity (30 to less than 40); class 3 i.e. severe obesity (40 or greater).

[h] Acute organ failure defined as a composite of respiratory failure (new receipt of high-flow nasal canula, non-invasive mechanical ventilation, or IMV), cardiovascular failure (use of vasopressors), or kidney failure (new receipt of kidney replacement therapy).

[i] A total of 182 patients (25.5%) were missing data on symptom onset date. The median time between symptom onset and hospital admission was calculated based on nonmissing data.

Abbreviations: RSV = respiratory syncytial virus; ICU = intensive care unit; BMI = body mass index; IMV = intensive mechanical ventilation.

**Supplementary Table 6. Prevalence of Underlying Conditions Associated with Respiratory Syncytial Virus Disease Among Bayesian Profiles[a] of 714 Patients Aged ≥60 Hospitalized with Respiratory Syncytial Virus — IVY Network, 26 Hospitals, January 2022–July 2024**

| | Overall (N=714) | 1 (N=460) | 2 (N=183) | 3 (N=52) | 4 (N=19) | P-value |
|---|---|---|---|---|---|---|
| **BMI category** | | | | | | 0.000121 |
| Underweight | 53 (7.4%) | 42 (9.1%) | 9 (4.9%) | 1 (1.9%) | 1 (5.3%) | |
| Healthy weight | 199 (27.9%) | 137 (29.8%) | 38 (20.8%) | 17 (32.7%) | 7 (36.8%) | |
| Overweight | 210 (29.4%) | 144 (31.3%) | 43 (23.5%) | 21 (40.4%) | 2 (10.5%) | |
| Obesity | 182 (25.5%) | 94 (20.4%) | 71 (38.8%) | 10 (19.2%) | 7 (36.8%) | |
| Severe obesity | 70 (9.8%) | 43 (9.3%) | 22 (12.0%) | 3 (5.8%) | 2 (10.5%) | |
| **Diabetes** | | | | | | <0.0001 |
| No | 460 (64.4%) | 354 (77.0%) | 54 (29.5%) | 43 (82.7%) | 9 (47.4%) | |
| Yes | 254 (35.6%) | 106 (23.0%) | 129 (70.5%) | 9 (17.3%) | 10 (52.6%) | |
| **Chronic kidney disease** | | | | | | <0.0001 |
| No | 519 (72.7%) | 427 (92.8%) | 33 (18.0%) | 42 (80.8%) | 17 (89.5%) | |
| Yes; ESRD | 49 (6.9%) | 1 (0.2%) | 48 (26.2%) | 0 (0%) | 0 (0%) | |
| Yes; without ESRD | 146 (20.4%) | 32 (7.0%) | 102 (55.7%) | 10 (19.2%) | 2 (10.5%) | |
| **Heart failure** | | | | | | <0.0001 |
| No | 497 (69.6%) | 387 (84.1%) | 62 (33.9%) | 38 (73.1%) | 10 (52.6%) | |
| Yes | 217 (30.4%) | 73 (15.9%) | 121 (66.1%) | 14 (26.9%) | 9 (47.4%) | |
| **Atherosclerotic CVD** | | | | | | <0.0001 |
| No | 552 (77.3%) | 389 (84.6%) | 109 (59.6%) | 41 (78.8%) | 13 (68.4%) | |
| Yes | 162 (22.7%) | 71 (15.4%) | 74 (40.4%) | 11 (21.2%) | 6 (31.6%) | |
| **Asthma** | | | | | | 0.0118 |
| No | 586 (82.1%) | 367 (79.8%) | 157 (85.8%) | 49 (94.2%) | 13 (68.4%) | |

| | Overall (N=714) | 1 (N=460) | 2 (N=183) | 3 (N=52) | 4 (N=19) | P-value |
|---|---|---|---|---|---|---|
| Yes | 128 (17.9%) | 93 (20.2%) | 26 (14.2%) | 3 (5.8%) | 6 (31.6%) | |
| **COPD** | | | | | | <0.0001 |
| No | 479 (67.1%) | 296 (64.3%) | 131 (71.6%) | 50 (96.2%) | 2 (10.5%) | |
| Yes; home O2 use | 61 (8.5%) | 40 (8.7%) | 4 (2.2%) | 0 (0%) | 17 (89.5%) | |
| Yes; no home O2 use | 174 (24.4%) | 124 (27.0%) | 48 (26.2%) | 2 (3.8%) | 0 (0%) | |
| **Other pulmonary** | | | | | | <0.0001 |
| No | 650 (91.0%) | 446 (97.0%) | 159 (86.9%) | 45 (86.5%) | 0 (0%) | |
| Yes; home O2 use | 24 (3.4%) | 2 (0.4%) | 0 (0%) | 3 (5.8%) | 19 (100%) | |
| Yes; no home O2 use | 40 (5.6%) | 12 (2.6%) | 24 (13.1%) | 4 (7.7%) | 0 (0%) | |
| **Transplant receipt** | | | | | | <0.0001 |
| No | 658 (92.2%) | 455 (98.9%) | 165 (90.2%) | 19 (36.5%) | 19 (100%) | |
| Yes | 56 (7.8%) | 5 (1.1%) | 18 (9.8%) | 33 (63.5%) | 0 (0%) | |
| **Hematalogic malignancy** | | | | | | <0.0001 |
| No | 676 (94.7%) | 460 (100%) | 183 (100%) | 15 (28.8%) | 18 (94.7%) | |
| Yes | 38 (5.3%) | 0 (0%) | 0 (0%) | 37 (71.2%) | 1 (5.3%) | |
| **Solid tumor** | | | | | | 0.000997 |
| No | 659 (92.3%) | 415 (90.2%) | 180 (98.4%) | 45 (86.5%) | 19 (100%) | |
| Yes | 55 (7.7%) | 45 (9.8%) | 3 (1.6%) | 7 (13.5%) | 0 (0%) | |
| **HIV** | | | | | | 0.598 |
| No | 703 (98.5%) | 451 (98.0%) | 181 (98.9%) | 52 (100%) | 19 (100%) | |
| Yes | 11 (1.5%) | 9 (2.0%) | 2 (1.1%) | 0 (0%) | 0 (0%) | |
| **Autoimmune or IBD** | | | | | | 0.0281 |
| No | 663 (92.9%) | 429 (93.3%) | 173 (94.5%) | 43 (82.7%) | 18 (94.7%) | |
| Yes | 51 (7.1%) | 31 (6.7%) | 10 (5.5%) | 9 (17.3%) | 1 (5.3%) | |
| **Dementia** | | | | | | 0.209 |

| | Overall (N=714) | 1 (N=460) | 2 (N=183) | 3 (N=52) | 4 (N=19) | P-value |
|---|---|---|---|---|---|---|
| No | 664 (93.0%) | 425 (92.4%) | 170 (92.9%) | 52 (100%) | 17 (89.5%) | |
| Yes | 50 (7.0%) | 35 (7.6%) | 13 (7.1%) | 0 (0%) | 2 (10.5%) | |
| **Neuromuscular disease** | | | | | | 0.924 |
| No | 705 (98.7%) | 454 (98.7%) | 181 (98.9%) | 51 (98.1%) | 19 (100%) | |
| Yes | 9 (1.3%) | 6 (1.3%) | 2 (1.1%) | 1 (1.9%) | 0 (0%) | |
| **Liver disease** | | | | | | 0.000108 |
| No | 683 (95.7%) | 450 (97.8%) | 168 (91.8%) | 46 (88.5%) | 19 (100%) | |
| Yes; cirrhosis | 24 (3.4%) | 10 (2.2%) | 11 (6.0%) | 3 (5.8%) | 0 (0%) | |
| Yes; without cirrhosis | 7 (1.0%) | 0 (0%) | 4 (2.2%) | 3 (5.8%) | 0 (0%) | |

Percentages are column percentages. *P*-values are calculated using the Chi-square test. Among adults aged ≥60, profile 1 represents the minimal prevalence profile, 2 represents the cardiorenal and diabetes profile, 3 represents the hematologic malignancy profile, and 4 represents the severe chronic pulmonary disease with home oxygen dependence profile.

[a] Bayesian profile regression is an outcome variable-informed statistical clustering approach that comprises an assignment submodel (assigns individuals to a condition profile) and a disease submodel (evaluates association of profiles with the outcome variable); the two submodels are jointly fitted in a Bayesian paradigm, enabling the outcome variable to influence profile membership.

Abbreviations: RSV = respiratory syncytial virus; BMI = body mass index.

**Supplementary Table 7. Association of Chronic Obstructive Pulmonary Disease and Acute Organ Failure[a] Among Bayesian Profiles[b] of 397 Patients Aged 18–59 Hospitalized with Respiratory Syncytial Virus — IVY Network, 26 Hospitals, January 2022–July 2024**

| Profile | COPD without home oxygen | Acute organ failure | N | Prevalence of outcome within profile and condition stratum (%) |
|---|---|---|---|---|
| **1** | No | No | 205 | 76.2 |
| **1** | No | Yes | 64 | 23.8 |
| **1** | Yes | No | 11 | 47.8 |
| **1** | Yes | Yes | 12 | 52.2 |
| **2** | No | No | 53 | 69.7 |
| **2** | No | Yes | 23 | 30.3 |
| **2** | Yes | No | 5 | 29.4 |
| **2** | Yes | Yes | 12 | 70.6 |

In this descriptive table, we assessed the association of the primary risk factor identified through Poisson regression among adults aged 18–59 (COPD without home oxygen) with the outcome of acute organ failure within levels of profiles identified using Bayesian profile regression. Profile 1 represents the minimal prevalence profile and 2 represents the cardiorenal and diabetes profile. The *P*-value from Fisher's exact test for the association between COPD without home oxygen use and acute organ failure among patients in profile 1 was 0.005 and among patients in profile 2 was 0.004.

[a] Acute organ failure was defined as a composite of respiratory failure (new receipt of high-flow nasal canula, non-invasive mechanical ventilation, or IMV), cardiovascular failure (use of vasopressors), or kidney failure (new receipt of kidney replacement therapy).

[b] Bayesian profile regression is an outcome variable-informed statistical clustering approach that comprises an assignment submodel (assigns individuals to a condition profile) and a disease submodel (evaluates association of profiles with the outcome variable); the two submodels are jointly fitted in a Bayesian paradigm, enabling the outcome variable to influence profile membership.

Abbreviations: COPD = chronic obstructive pulmonary disease; IMV = invasive mechanical ventilation.

**Supplementary Figure 1. Numbers of Underlying Conditions[a] Associated with Respiratory Syncytial Virus Disease by 10-Year Age Bands Among Patients Hospitalized with Respiratory Syncytial Virus — IVY Network, 26 Hospitals, January 2022–July 2024**

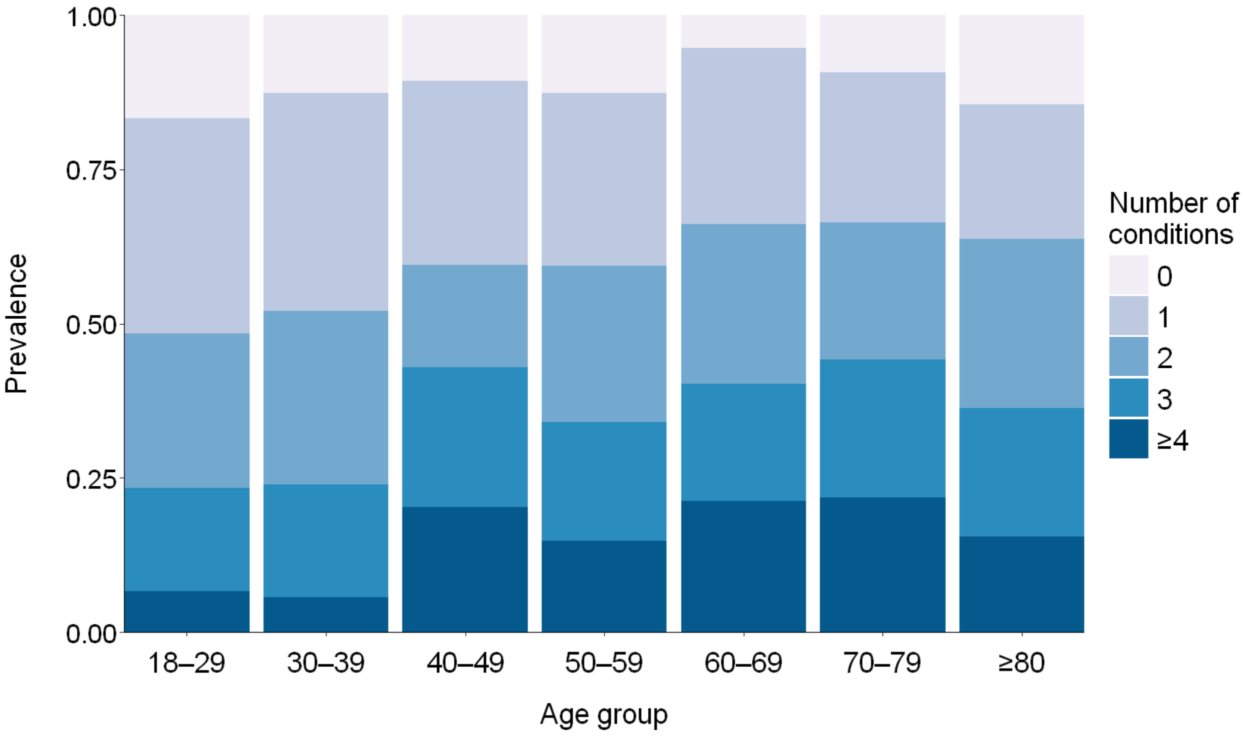

[a] BMI categories were defined using the following BMI (kg/m$^2$) ranges: underweight (less than 18.5); healthy weight (18.5 to less than 25); overweight (25 to less than 30); class 1 or 2 obesity (30 to less than 40); class 3 i.e. severe obesity (40 or greater). For counts of underlying conditions, only severe obesity was included among all BMI categories to align with vaccine eligibility guidelines [1]. Other underlying conditions associated with RSV severe disease and included in counts were diabetes, chronic kidney disease (including end-stage renal disease), heart failure, atherosclerotic cardiovascular disease, asthma, chronic obstructive pulmonary disease (with or without home oxygen use), other pulmonary conditions (cystic fibrosis, pulmonary fibrosis, or pulmonary hypertension; with or without home oxygen use), solid organ or hematopoietic cell transplant receipt, active hematologic malignancy, active solid tumor malignancy, HIV infection, autoimmune or autoinflammatory conditions (systemic lupus erythematosus, rheumatoid arthritis, psoriasis, polymyositis, mixed connective tissue disease, polymyalgia rheumatica, inflammatory bowel disease, or other autoimmune diseases), dementia (only included for adults aged ≥60), neuromuscular conditions (muscular dystrophy, cerebral

palsy, hemiplegia, paraplegia, or anterolateral sclerosis), liver disease (including cirrhosis), and sickle cell disease or thalassemia (only included for adults aged 18–59).

**Supplementary Figure 2. Pairwise Correlation of Underlying Conditions[a] Among Patients Hospitalized with Respiratory Syncytial Virus — IVY Network, 26 Hospitals, January 2022–July 2024**

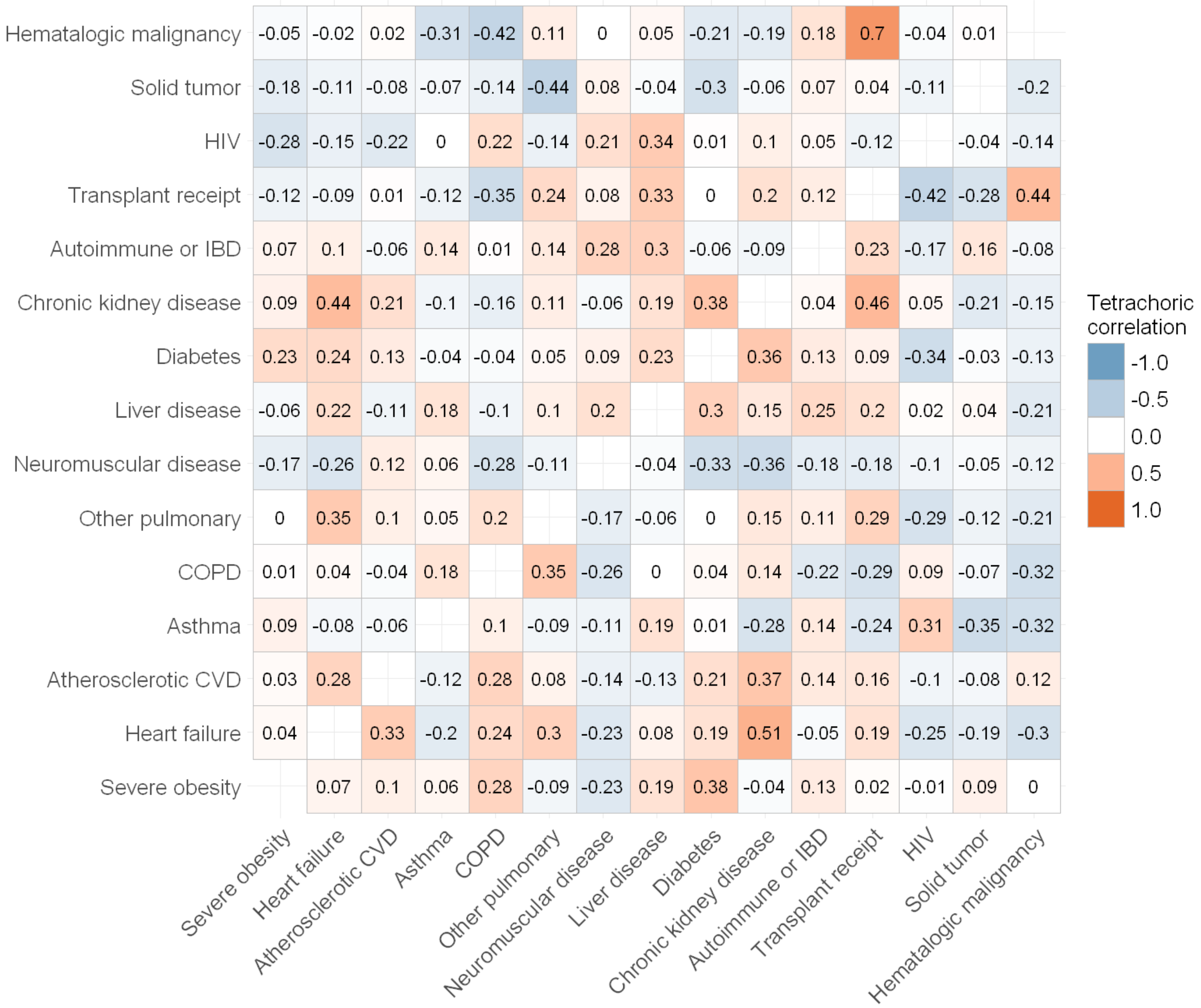

Correlation among adults 18–59 is shown in the lower triangular matrix, and correlation among adults ≥60 is shown in the upper triangular matrix.

[a] Only underlying conditions with prevalence 1% or greater in both age groups were included. Among the BMI categories, only severe obesity was included. Chronic obstructive pulmonary disease and other pulmonary conditions were included without stratifying by home oxygen use. Other pulmonary conditions included cystic fibrosis, pulmonary fibrosis, or pulmonary hypertension. Neuromuscular conditions included muscular dystrophy, cerebral palsy, hemiplegia, paraplegia, or anterolateral sclerosis. Autoimmune or autoinflammatory conditions

included systemic lupus erythematosus, rheumatoid arthritis, psoriasis, polymyositis, mixed connective tissue disease, polymyalgia rheumatica, inflammatory bowel disease, or other autoimmune diseases.

**Supplementary Figure 3. Age Distribution Among Bayesian Profiles[a] of Patients Hospitalized with Respiratory Syncytial Virus — IVY Network, 26 Hospitals, January 2022–July 2024**

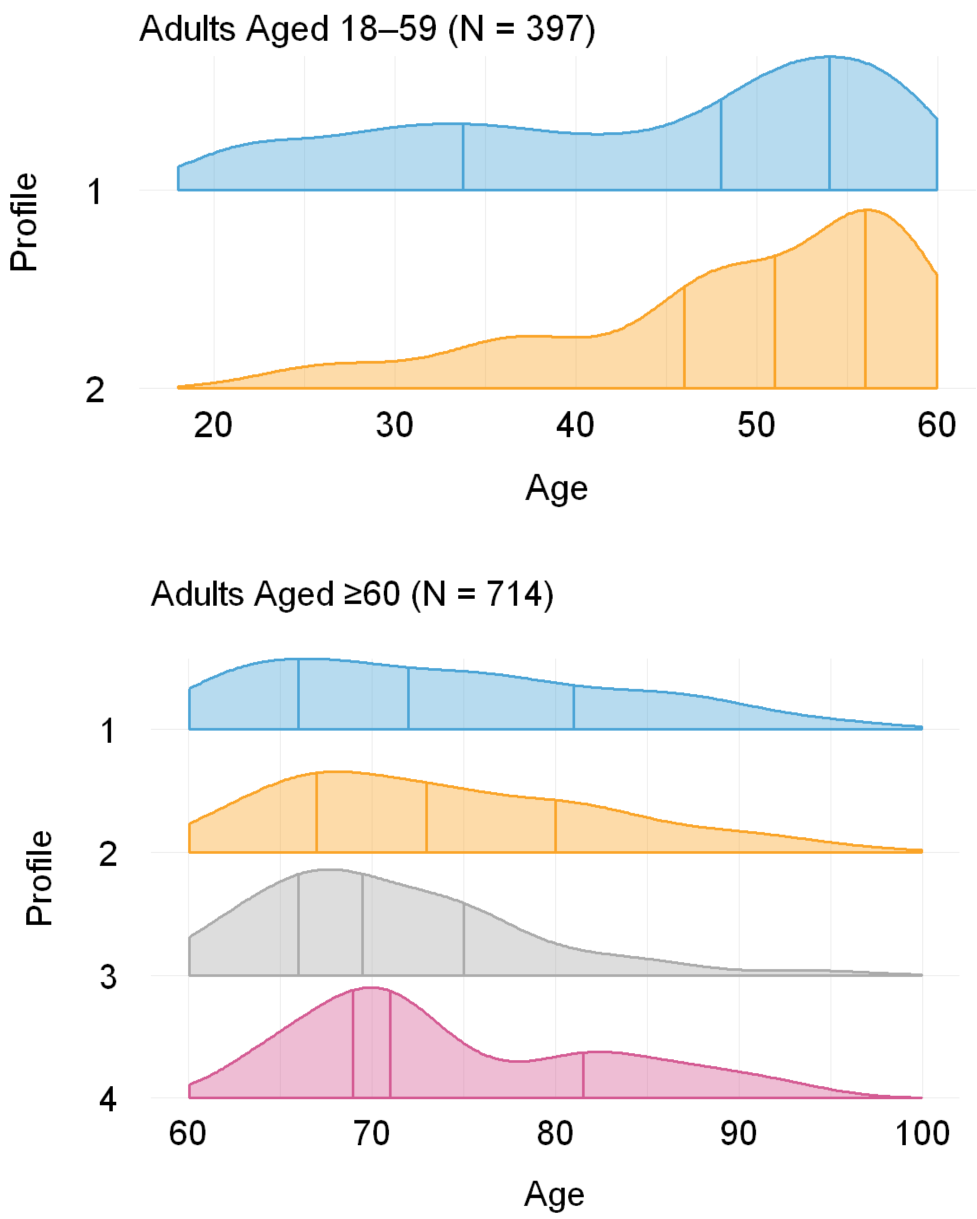

First quartiles, medians, and third quartiles are indicated by the first, second, and third vertical lines within a distribution, respectively. Among adults in both age groups, profile 1 represents the minimal prevalence profile and 2 represents the cardiorenal and diabetes profile. Among adults aged ≥60, profile 3 represents the hematologic malignancy profile, and 4 represents the severe chronic pulmonary disease with home oxygen dependence profile.

[a] Bayesian profile regression is an outcome variable-informed statistical clustering approach that comprises an assignment submodel (assigns individuals to a condition profile) and a disease submodel (evaluates association of profiles with the outcome variable); the two submodels are jointly fitted in a Bayesian paradigm, enabling the outcome variable to influence profile membership.

**Supplementary Figure 4. Risk and 95% Confidence Intervals of Severe In-Hospital Outcome[a] by Condition[a] Among 397 Patients Aged 18–59 Hospitalized with Respiratory Syncytial Virus Estimated Using Poisson Regression[c] — IVY Network, 26 Hospitals, January 2022–July 2024**

(Figure on following page)

Risk estimates are presented with 95% confidence intervals unadjusted for multiple testing. Conditions where the 95% confidence interval exclude the null correspond to those indicated with a '*' in main Figure 4.

[a] Severe outcomes included ICU admission, acute organ failure, and invasive mechanical ventilation (IMV) or death. Acute organ failure was defined as a composite of respiratory failure (new receipt of high-flow nasal canula, non-invasive mechanical ventilation, or IMV), cardiovascular failure (use of vasopressors), or kidney failure (new receipt of kidney replacement therapy).

[b] Underlying conditions potentially associated with RSV disease included underweight, overweight, class 1 or 2 obesity, severe obesity, diabetes, chronic kidney disease (including end-stage renal disease), heart failure, atherosclerotic cardiovascular disease, asthma, chronic obstructive pulmonary disease (with or without home oxygen use), other pulmonary conditions (cystic fibrosis, pulmonary fibrosis, or pulmonary hypertension; with or without home oxygen use), solid organ or hematopoietic cell transplant receipt, active hematologic malignancy, active solid tumor malignancy, HIV infection, autoimmune or autoinflammatory conditions (systemic lupus erythematosus, rheumatoid arthritis, psoriasis, polymyositis, mixed connective tissue disease, polymyalgia rheumatica, inflammatory bowel disease, or other autoimmune diseases), dementia (only included for adults aged ≥60), neuromuscular conditions (muscular dystrophy, cerebral palsy, hemiplegia, paraplegia, or anterolateral sclerosis), liver disease (including cirrhosis), and sickle cell disease or thalassemia (only included for adults aged 18–59).

[c] To estimate risk ratios while accounting for rare conditions, we fit multivariable Firth bias-reduced Poisson regression models using robust error variances with the Morel correction using the *firthb* R package.

Abbreviations: RSV = respiratory syncytial virus; ICU = intensive care unit; IMV = intensive mechanical ventilation; BMI = body mass index; CVD = cardiovascular disease; COPD = chronic obstructive pulmonary disease; IBD = inflammatory bowel disease.

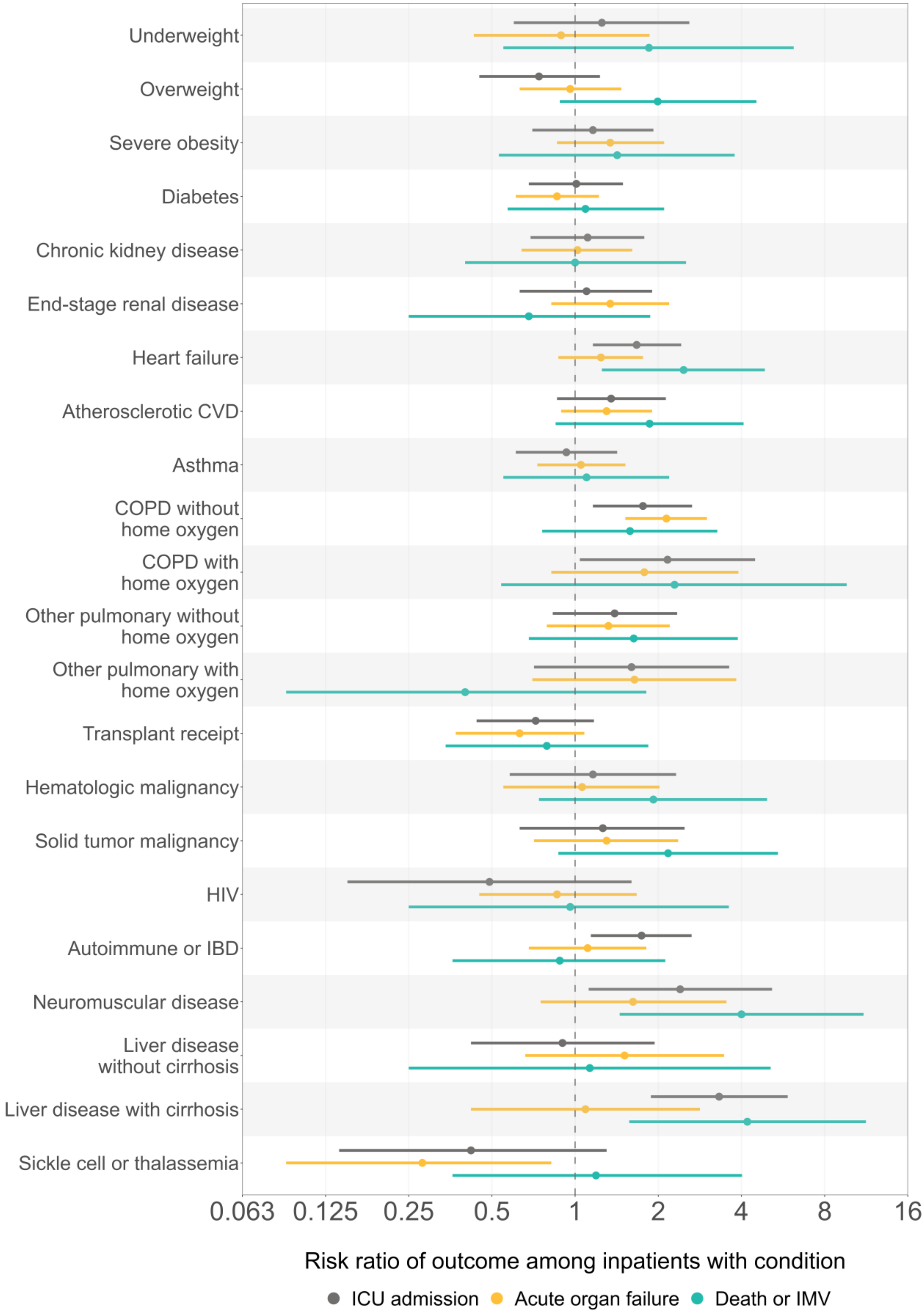
Underweight
Overweight
Severe obesity
Diabetes
Chronic kidney disease
End-stage renal disease
Heart failure
Atherosclerotic CVD
Asthma
COPD without home oxygen
COPD with home oxygen
Other pulmonary without home oxygen
Other pulmonary with home oxygen
Transplant receipt
Hematologic malignancy
Solid tumor malignancy
HIV
Autoimmune or IBD
Neuromuscular disease
Liver disease without cirrhosis
Liver disease with cirrhosis
Sickle cell or thalassemia
0.063
0.125
0.25
0.5
1
2
4
8
16
Risk ratio of outcome among inpatients with condition
ICU admission
Acute organ failure
Death or IMV

**Supplementary Figure 5. Risk and 95% Confidence Intervals of Severe In-Hospital Outcome[a] by Condition[b] Among 714 Patients Aged ≥60 Hospitalized with Respiratory Syncytial Virus Estimated Using Poisson Regression[c] — IVY Network, 26 Hospitals, January 2022–July 2024**

(Figure on following page)

Risk estimates are presented with 95% confidence intervals unadjusted for multiple testing. Conditions where the 95% confidence interval exclude the null correspond to those indicated with a '*' in main Figure 4.

[a] Severe outcomes included ICU admission, acute organ failure, and invasive mechanical ventilation (IMV) or death. Acute organ failure was defined as a composite of respiratory failure (new receipt of high-flow nasal canula, non-invasive mechanical ventilation, or IMV), cardiovascular failure (use of vasopressors), or kidney failure (new receipt of kidney replacement therapy).

[b] Underlying conditions potentially associated with RSV disease included underweight, overweight, class 1 or 2 obesity, severe obesity, diabetes, chronic kidney disease (including end-stage renal disease), heart failure, atherosclerotic cardiovascular disease, asthma, chronic obstructive pulmonary disease (with or without home oxygen use), other pulmonary conditions (cystic fibrosis, pulmonary fibrosis, or pulmonary hypertension; with or without home oxygen use), solid organ or hematopoietic cell transplant receipt, active hematologic malignancy, active solid tumor malignancy, HIV infection, autoimmune or autoinflammatory conditions (systemic lupus erythematosus, rheumatoid arthritis, psoriasis, polymyositis, mixed connective tissue disease, polymyalgia rheumatica, inflammatory bowel disease, or other autoimmune diseases), dementia (only included for adults aged ≥60), neuromuscular conditions (muscular dystrophy, cerebral palsy, hemiplegia, paraplegia, or anterolateral sclerosis), liver disease (including cirrhosis), and sickle cell disease or thalassemia (only included for adults aged 18–59).

[c] To estimate risk ratios while accounting for rare conditions, we fit multivariable Firth bias-reduced Poisson regression models using robust error variances with the Morel correction using the *firthb* R package.

Abbreviations: RSV = respiratory syncytial virus; ICU = intensive care unit; IMV = intensive mechanical ventilation; BMI = body mass index; CVD = cardiovascular disease; COPD = chronic obstructive pulmonary disease; IBD = inflammatory bowel disease.

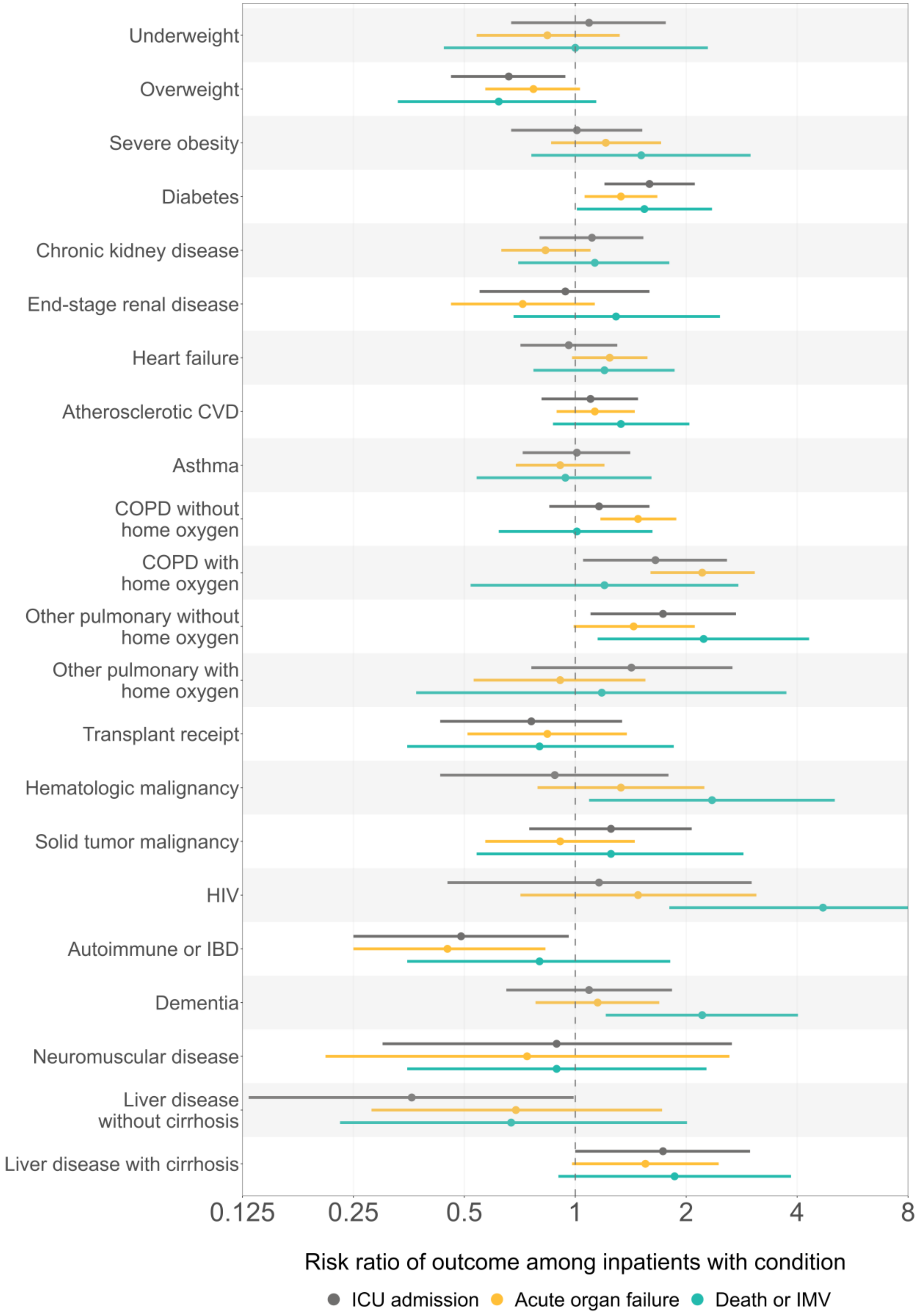
Underweight
Overweight
Severe obesity
Diabetes
Chronic kidney disease
End-stage renal disease
Heart failure
Atherosclerotic CVD
Asthma
COPD without home oxygen
COPD with home oxygen
Other pulmonary without home oxygen
Other pulmonary with home oxygen
Transplant receipt
Hematologic malignancy
Solid tumor malignancy
HIV
Autoimmune or IBD
Dementia
Neuromuscular disease
Liver disease without cirrhosis
Liver disease with cirrhosis
0.125
0.25
0.5
1
2
4
8
Risk ratio of outcome among inpatients with condition
ICU admission
Acute organ failure
Death or IMV
